# Genome-wide association study of obstructive sleep apnea in the Million Veteran Program uncovers genetic heterogeneity by sex

**DOI:** 10.1101/2022.12.21.22283799

**Authors:** Tamar Sofer, Nuzulul Kurniansyah, Michael Murray, Yuk-Lam Ho, Jennifer E. Huffman, Kelly Cho, Peter W.F. Wilson, Daniel J Gottlieb, the VA Million Veteran Program

**Affiliations:** Division of Sleep and Circadian Disorders, Department of Medicine, Brigham and Women’s Hospital, Boston, MA; Department of Biostatistics, Harvard T.H. Chan School of Public Health, Boston, MA; Massachusetts Veterans Epidemiology Research and Information Center, Veterans Affairs Boston Healthcare System, Boston, MA; VA Palo Alto Health Care System, Palo Alto, CA, USA; Palo Alto Veterans Institute for Research, Palo Alto, CA, USA; Atlanta VA Healthcare System, Decatur, GA

## Abstract

**Background:** Genome-wide association studies (GWAS) for obstructive sleep apnea (OSA) are limited due to the underdiagnosis of OSA, leading to misclassification of OSA, which consequently reduces statistical power. We performed a GWAS of OSA in the Million Veteran Program (MVP) of the U.S. Department of Veterans Affairs (VA) healthcare system, where OSA prevalence is close to its true population prevalence.

**Methods:** We performed GWAS of 568,576 MVP participants, stratified by biological sex and by harmonized race/ethnicity and genetic ancestry (HARE) groups of White, Black, Hispanic, and Asian individuals. We considered both BMI adjusted (BMI-adj) and unadjusted (BMI-unadj) models. We replicated associations in independent datasets, and analyzed the heterogeneity of OSA genetic associations across HARE and sex groups. We finally performed a larger meta-analysis GWAS of MVP, FinnGen, and the MGB Biobank, totaling 916,696 individuals.

**Findings:** MVP participants are 91% male. OSA prevalence is 21%. In MVP there were 18 and 6 genome-wide significant loci in BMI-unadj and BMI-adj analyses, respectively, corresponding to 21 association regions. Of these, 17 were not previously reported in association with OSA, and 13 replicated in FinnGen (False Discovery Rate p-value<0.05). There were widespread significant differences in genetic effects between men and women, but less so across HARE groups. Meta-analysis of MVP, FinnGen, and MGB biobank revealed 17 additional, novel, genome-wide significant regions.

**Interpretation:** Sex differences in genetic associations with OSA are widespread, likely associated with multiple OSA risk factors. OSA shares genetic underpinnings with several sleep phenotypes, suggesting shared etiology and causal pathways.

**Funding:** Described in acknowledgements.

## Introduction

Obstructive sleep apnea (OSA) is a condition characterized by repeated airway collapses during sleep, leading to intermittent reductions in oxyhemoglobin saturation and cortical arousals during sleep (1). Much like other health phenotypes, OSA and its related quantitative phenotypes such as the apnea-hypopnea index (AHI) have a genetic basis. The heritabilities of OSA-related phenotypes have been estimated to be up to 25%-40% in family studies (2–4), and up to 17% in population-based studies (5,6). Genome-wide association studies (GWAS) of OSA and related phenotypes reported a handful of genetic variants (6–11), mostly related to cardiometabolic conditions such as obesity and lung function. There are known sex, gender and race/ethnic differences in OSA (12–14), but their genetic basis has not been thoroughly studied, mainly due to sample size and resulting power limitations. Because BMI and other adiposity and body fat distribution measures are strong causal determinants of OSA (15), it is important to account for it when studying the genetic basis of OSA. Until recently, the largest GWAS of OSA and of a proxy phenotype, snoring, primarily reported results from analyses that did not adjust for BMI (16,17). A recent publication made a considerable progress and reported a multi-trait analysis of OSA and snoring, followed with replication analysis of top associations in a BMI-adjusted analysis in 23andMe (18).

Critically for GWAS, OSA is common yet under-diagnosed (19,20). In healthcare settings, OSA is typically evaluated by indication, following symptoms such as excessive daytime sleepiness and in individuals who are perceived to be more likely to have OSA, i.e., those who are overweight or obese (21). Thus, individuals with OSA are often not diagnosed. For example, a publication based on health records reported <1% individuals with the diagnosis of OSA (22), while another study identified 9% prevalence of moderate-to-severe OSA among individuals without OSA history (23). Estimates from population-based cohort studies in adults, which are not biased by indication for assessment, are 9-38% for mild to severe OSA and 2-17% of moderate to severe OSA (24,25). Thus, statistical power for genetic analysis of OSA is often limited: while OSA can be studied based on large biobanks, its underdiagnosis limits this investigation: e.g., only 1.47% of UK Biobank individuals have OSA according to the electronic health records (17). In FinnGen, based on which a GWAS of OSA was recently published, the OSA rate was 8% (6), which also likely points to misclassification of many “controls”. In the 23andMe analysis reported by Campos et al. (18), ∼12% participants self-reported OSA. Population-based cohort studies which measure all individuals regardless of indications are available, and were used for GWAS of OSA-related measures, but their sample sizes are limited (9–11,26).

In contrast to the prevalence of OSA identified from health records in other healthcare systems, a high prevalence of OSA has been identified in patients receiving care from the U.S. Department of Veterans Affairs (VA) healthcare system, with a reported prevalence of sleep-related breathing disorders of 22.2% in 2018, based on International Classification of Disease (ICD) codes (27). This reflects a 4-fold increase in prevalence from 2012 and follows a 6-fold increase between 2000 and 2010 (28). While the prevalence of OSA in the VA patient population is likely to exceed that in the general population, other factors have been largely responsible for the increase in identification of OSA, including the early and widespread adoption of home sleep apnea testing in the VA, the low cost to Veterans for diagnostic services, the lack of copayments for prosthetic equipment used in OSA treatment, and financial incentives for service-related illnesses providing an incentive for patients with sleep problems to pursue a diagnosis. The prevalence of diagnosed OSA in the VA is therefore much closer to the expected population prevalence of OSA when compared to reports from other healthcare settings, likely resulting in lower misclassification rate (i.e., fewer individuals with OSA being identified as not having OSA in the medical record). Here, data from 568,576 individuals from the VA’s Million Veteran Program (MVP) contributed to the GWAS of OSA. Analyses accounted for BMI and stratified by sex and by groups approximating race and ethnicity, while prioritizing genetic similarity. We investigated genetic determinants of OSA and their heterogeneity across these population subgroups, and genetic relationship to other phenotypes. We also performed a larger meta-analysis GWAS of MVP, FinnGen, and the Mass General Brigham (MGB) Biobank, totaling 916,696 individuals.

## Methods

### Obstructive sleep apnea phenotype definition and covariates

All participants were Veterans and OSA was identified from the VA electronic health record using the previously published multimodal automated phenotyping (MAP) procedure to provide a predicted probability that a patient has the phenotype of interest and a threshold probability for a binary classification of the phenotype as present or absent (29). MAP is an unsupervised clustering algorithm that uses counts of ICD codes from the electronic medical record and concept unique identifiers (CUI) derived by natural language processing of text notes, along with total number of notes as a proxy for healthcare utilization. For this analysis, the MAP procedure utilized ICD-9 code 327.23 for OSA, 327.2x, 780.51, 780.53, 780.57 for SA, ICD-10 code G47.33 for OSA, G47.3X for SA and CUI C0520679 (Sleep Apnea, Obstructive) for OSA, and C1561861, C1561869, C0037315, C0751762, C1561862 for SA in the Unified Medical Language System (https://uts.nlm.nih.gov/uts/umls/concept/C0520679). The MAP probability cutoffs were 0.38 for MAP-OSA and 0.37 for MAP-SA. The OSA phenotype for this analysis included all individuals with MAP-OSA ≥0.38 plus those with both a MAP-OSA >0 and MAP-SA ≥0.37. This phenotype was validated by reviewing 100 charts randomly selected from patients who had at least one ICD code for OSA, and it had a positive predictive value of 0.92.

Analyses were adjusted for age at the first OSA ICD code in OSA cases and at the age during last VA encounter in OSA controls, and the first 10 PCs of genetic data. In addition, a BMI adjusted model further adjusted for BMI, using both linear and squared terms. BMI measures were taken from the electronic health records from a visit within a year of the participant’s MVP enrollment.

### Genotyping and imputation

Genotyping, imputation, and relatedness inference were previously described (30,31). Briefly, study participants were genotyped using a customized Affymetrix Axiom Biobank Array (31,32). Imputation was performed to a hybrid imputation panel comprised of the African Genome Resources panel (https://imputation.sanger.ac.uk/?about=1#referencepanels) and 1000 Genomes (p3v5). Principal components (PCs) of genetic data were computed using EIGENSOFT v.6 (33) based on genotyped variants. The harmonized race/ethnicity and genetic ancestry (HARE) approach was used to assign individuals to four groups: non-Hispanic White (White), non-Hispanic Black (Black), Hispanic or Latino (Hispanic), and Asian (34). Kinship coefficients were inferred using KING v.2.0 (35). We remove related individuals with kinship coefficient ≥0.0884, so that one individual was excluded from each related pair, preferentially retaining those who have OSA. All analyses used the unrelated set of individuals. Imputation quality scores (INFO) were computed separately in each HARE group.

When testing variant associations within any HARE group, we excluded variants with imputation quality INFO<0.3 and with MAF<0.01 within this group. Final index SNPs representing any reported association regions had to have imputation quality INFO≥0.8 in the combined sample.

### Genome-wide association study

We performed association analyses using PLINK v2.00a3LM (36), separately by HARE group and biological sex (confirmed by sex chromosomes), followed by meta-analysis using GWAMA v2.2.2 (37) with genomic control. As secondary findings, we also report results from analyses by HARE and by sex groups, and a multi-population meta-analysis stratified by obesity, and use them to study heterogeneity in associations across strata (more below).

Because multiple independent associations may be obtained in the same genomic region, we performed clumping using the relevant dataset as the linkage disequilibrium (LD) reference panel (e.g., for findings in the primary, sex-combined, multi-population meta-analysis, we used all individuals). Clumping was performed using PLINK v1.9 with clumping parameters R^2^=0.1 and distance =1000Kbp. SNPs were annotated to their nearest gene using the FAVOR variant annotation database (38). Regional association plots were generated for each of the top SNPs after clumping, using 1Mbp regions centered at each SNP. Genes positions for the regional association plots were extracted from the UCSC genome browser (39). We looked up previously reported associations with OSA SNPs by a manual search in the Type 2 Diabetes Knowledge Portal (https://t2d.hugeamp.org/). We determined that a SNP was not previously reported as associated with another sleep phenotypes if it was not associated with any sleep disordered breathing phenotypes (including snoring, apnea-hypopnea index, and oxyhemoglobin saturation phenotypes), self-reported sleep phenotypes (e.g. naps, sleep duration, insomnia), or actigraphy-based sleep phenotypes at the 5×10^−8^ level. Summary statistics from GWAS of these phenotypes are available on the Type 2 Diabetes Knowledge Portal.

### Replication analysis

To test the detected associations for replication, we looked up top variants in publicly available GWAS of Finnish White individuals from FinnGen version 7 (N=307,927: 27,207 cases and 280,720 controls). Summary statistics were downloaded from the FinnGen portal. Because the FinnGen OSA GWAS was not adjusted for BMI, we also downloaded summary statistics from a FinnGen GWAS of obesity. Then, we performed an approximated conditional analysis using the mt-COJO procedure implemented in GCTA v1.93.2 (40). This resulted in GWAS summary statistics for OSA adjusted for obesity status. Because mt-COJO requires an LD reference panel, we used the 1000 Genomes Europeans (CEU) as well as the precomputed LD scores based on the CEU population. An association was deemed replicated if its one-sided p-value, with direction determined by the discovery direction of association (41), was <0.05. We also report false discovery rate (FDR) adjusted replication p-values, computed using the Benjamini-Hochberg procedure (42) over all primary multi-population BMI-adj and BMI-unadj associations. We also performed association look-ups in a GWAS of snoring from the UK Biobank (17).

### Genetic risk scores using top associated variants

In MGB Biobank, we constructed genetic risk scores for OSA based on the independent genetic variants identified in the multi-population analysis. We constructed a few genetic risk scores: unweighted, a sum of risk-increasing alleles (GRS); weighted score, where risk alleles were weighted by their estimated effect sizes (wGRS); and both based on multi-population BMI-adj and BMI-unadj analyses. We report the associations of the GRS and wGRS in the MGB biobank stratified by sex, and in sex-combined analysis. In addition, we estimated all associations in MGB biobank both BMI-adj and BMI-unadj analyses. The (w)GRS is modelled as continuous, with effect size estimated per 1 standard deviation increase in the (w)GRS.

### Analysis of the heterogeneity of genetic associations with OSA across genetic background and sex

We analyzed the evidence for genetic heterogeneity of OSA across genetic background and sex, in accordance with the known phenotypic heterogeneity. First, we estimated the heritability of OSA across HARE and biological sex groups. Heritability estimation is described below. Next, we tested differences in genetic associations between males and females, and between Black and White HARE groups (sex combined). Because all individuals were unrelated, we used as a test statistic for the *jth*genetic variant:

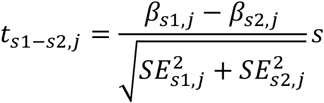

Where *β*_*s*1,*j*_, *β*_*s*2,*j*_ are the estimated log-OR of the rare allele of the variants in strata *s*1, *s*2 and *SE*_*s*1,*j*_, *SE*_*s*2,*j*_ are the corresponding estimated SEs. We annotated genome-wide significant sex-different and HARE-different SNPs by manual extraction of associations with p-value<0.001 from the Type 2 Diabetes Knowledge Portal.

We also evaluated the evidence of associations generalizing across discovery strata. For each sex, HARE, and obesity stratum, as well as the multi-population analysis, we identified SNP genome-wide significant associations, and evaluated the evidence for their associations in other strata. Thus, we tabulated how many of the SNPs had p-value<5×10^−8^ or <0.05 in other strata. In addition, to account for differences in power due to sample sizes and allele frequencies, we followed (43) and computed the power for identifying associations in the given stratum at the p-value<0.05 level, and, for each SNP category defined by discovery GWAS (e.g. multi-population, male, Hispanic, etc.), we summed the estimated power to produce the expected number of generalized associations.

### Meta-analysis with FinnGen and MGB Biobank

In a composite analysis, we meta-analyzed the GWAS of multi-population individuals from MVP, the FinnGen OSA GWAS, and GWAS of (mostly White) unrelated individuals from the Mass General Brigham (MGB) Biobank (N BMI-unadj = 40,193, N BMI-adj = 35,960) using GWAMA v2.2.2 (37). We performed clumping (1000Kbp, R^2^=0.1) to define the top hits using MVP multi-population genotypes as a reference panel.

### Heritability estimation

We calculated population-specific LD scores using LDSC (44) based on MVP participants for White, Black, Hispanic, and Asian HARE groups, and for the combined sample, using SNPs selected from HapMap 3 (45). We excluded SNPs from the major histocompatibility complex (MHC) region (chr6: 26–34Mb; grch37), SNPs with imputation INFO ≤0.9 and MAF ≤ 0.01 in each group. SNP-based heritability was estimated from the MVP GWAS summary statistics within HARE groups and in the multi-population dataset, by sex (male, female, and everyone), with and without BMI adjustment, and stratified by obesity (sex combined). For small groups (<25,000 individuals), including only females in each of the HARE groups, and the Asian HARE groups (female, male, and everyone) we computed the heritability using individual-level data using GCTA v.1.93.2.

### Genetic correlation estimation

To assess the impact of BMI on the genetic basis of OSA, we estimated genetic correlation using LDSC (46) between OSA “phenotypes” defined by BMI-adj and BMI-unadj analyses. Thus, in each HARE group, we estimated genetic correlation between the estimated genetic effects based on different analyses of OSA: BMI-adj, BMI-unadj, and within obesity groups (not adjusted for BMI). We excluded the Asian HARE group due to the small sample size.

We also estimated SNP-based genetic correlation between OSA and a range of additional phenotypes from sleep, cardiovascular, glycemic, psychiatric, and cognitive domains in sex-combined multi-population analyses. We studied the extent to which genetic correlations are attributable to the impact of BMI on OSA and the other phenotypes, using summary statistics that were either adjusted or unadjusted for BMI. To do this, we performed GWAS of each of the phenotypes in the MVP dataset using a smaller set of >800,000 SNPs – the HapMap 3 SNPs.

### Association of top SNPs with other phenotypes – assessment of pleiotropy

We tested the association of the top OSA-associated SNPs in the multi-population analysis with a range of cardiovascular, metabolic, and psychologic outcomes in MVP. The outcome definitions and adjusting covariates are described in the **Supplementary Information**.

### Gene-based analyses

We performed gene-based association analysis using the Multi-marker Analysis of GenoMic Annotation (MAGMA) software v1.10 (47). We used summary statistics from multi-population sex-combined and sex-specific analyses, and based on both BMI-unadj and BMI-adj model. We assigned SNPs to 19,427 protein coding genes, with gene locations provided in the MAGMA’s website (https://ctg.cncr.nl/software/magma, file: Gene locations, build 37). We followed others (48) and assigned SNPs to genes if they were within the window defined by 35kb downstream of the gene end position and 10kb upstream of the gene start positions. We used the SNP-wise model and randomly selected 10,000 unrelated multi-population MVP individuals to form a reference panel.

## Results

Data from N=568,576 contributed to the primary MVP dataset, with an overall OSA prevalence of 21.3%. Participants were 64 years old on average (standard deviation 15 years). 91.3% of the participants are male. **Figure 1** visualizes the prevalence of OSA in the MVP dataset, and the distribution of BMI, stratified by HARE and biological sex groups. OSA is most prevalent in the Hispanic male group (25.6%), and least common in the Asian female group (11.9%). **Supplemental Table 1** characterize the MVP study population by HARE group and combined. We performed genetic analysis of OSA according to the flowchart depicted in **Figure 2**.

**Figure 1:**
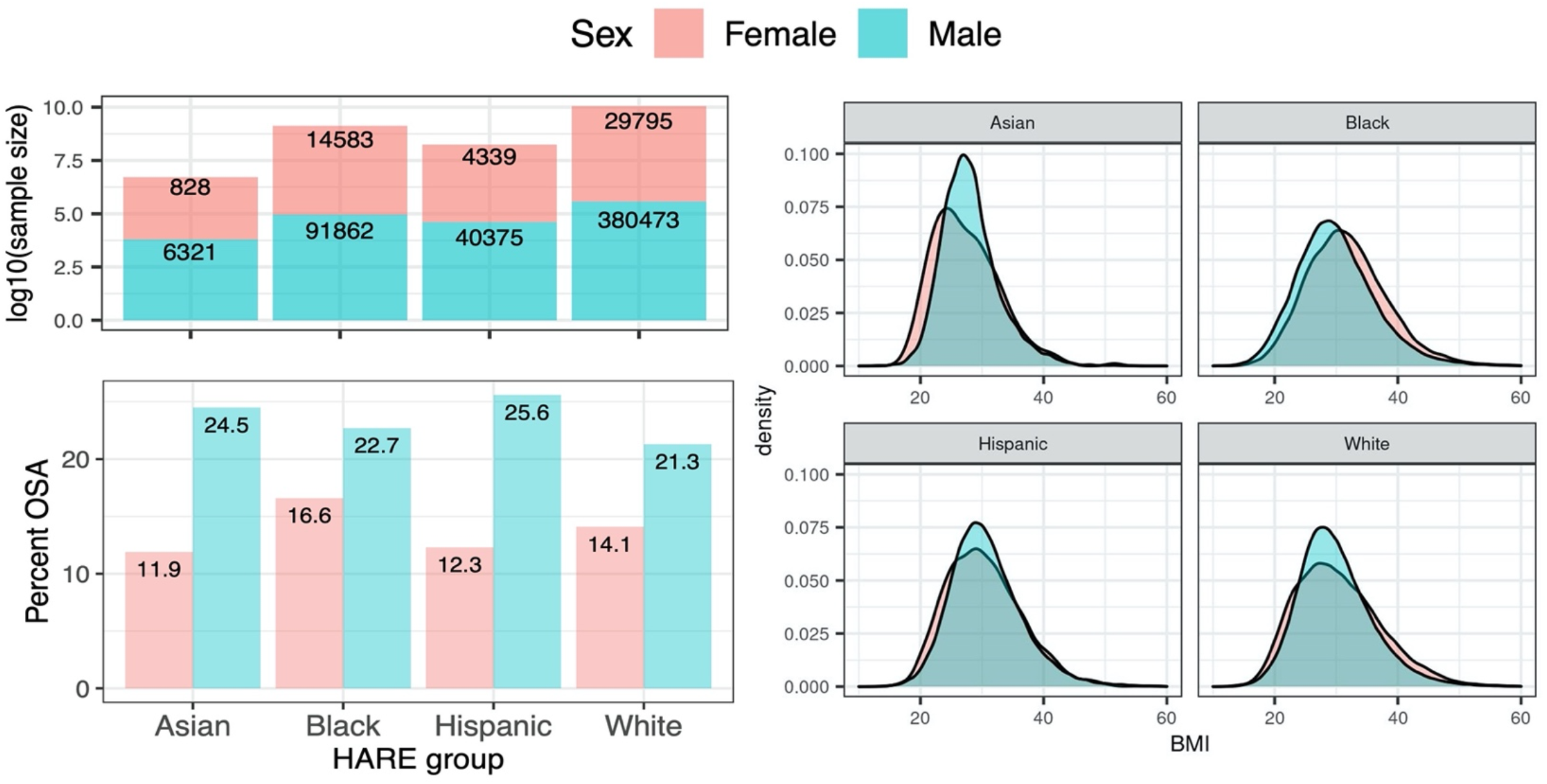
Proportions of OSA and BMI distribution across MVP HARE groups. Top left: number of OSA participants by HARE group and biological sex. Bottom left: Proportion of MVP participants who have OSA according to the EHR, stratified by HARE group and by sex. Right: density plots of BMI stratified by HARE and sex.

**Figure 2.**
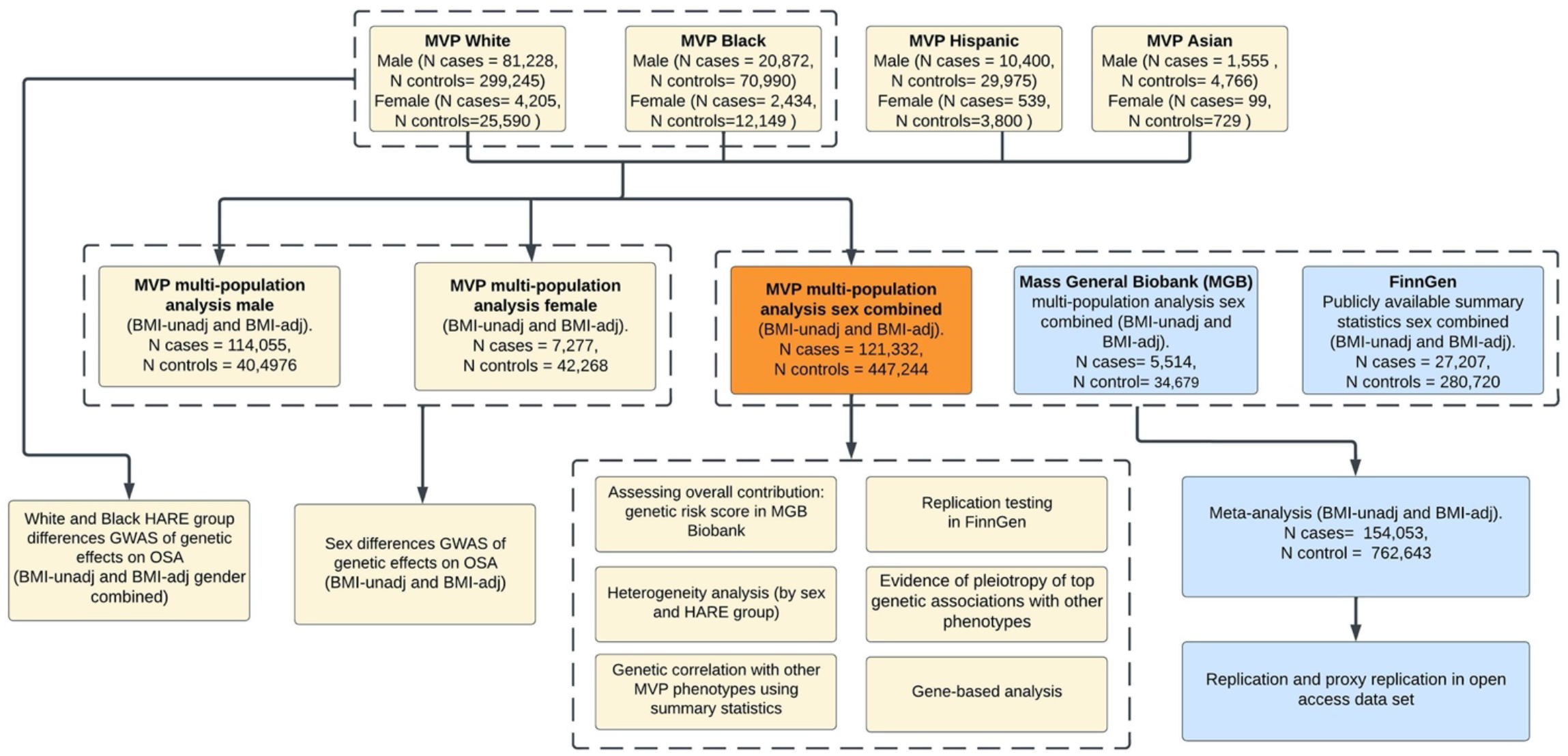
Flowchart of genetic analyses of OSA. Flowchart of genetic analyses of OSA performed. The primary analysis is the OSA GWAS in MVP. Various analyses were performed using the primary results. The MVP OSA GWAS summary statistics were also meta-analyzed with GWAS summary statistics from MGB Biobank and FinnGen, followed by replication and proxy-replication look up in open-access datasets.

### Primary GWAS and replication

**Figure 3 panel a** provides a Miami plot visualizing the findings and annotating the nearest gene for each association region. QQ-plots are provided in **Supplementary Figure 1** and regional association plots are provided in **Supplementary Figures 2** (BMI-unadj) and **3** (BMI-adj). In the primary GWAS, there were 18 genome-wide significant association regions in the BMI-unadj analysis, and 6 in the BMI-adj analysis (**Table 1**). Of these, from the BMI-unadj analysis, 2 associations were not available in FinnGen (nor other genome-wide significant SNPs from the same regions), and 13 associations replicated (one-sided p-value<0.05). Notably, all associations had the same direction of associations in MVP and in FinnGen. Of the 6 associations identified in BMI-adj analysis, 4 replicated in FinnGen and only one association had opposite direction of association between MVP and FinnGen. (**Table 1**). Of the replicated associations (BMI-unadj and BMI-adj), 8 associations were not previously reported for any sleep phenotype at the genome-wide significance level, and 9 were previously reported as associated with one or more sleep phenotype (snoring, sleep duration, napping, etc.). **Supplementary Table 2** reports previously reported associations of SNPs underlying genome-wide significant associations of OSA from **Table 1** with sleep traits, and summarizes the evidence of replication of associations.

**Table 1.** Top variants associated with OSA in multi-ethnic analysis (sex combined) in BMI-adj and -unadj analyses.

|  |  |  |  |  |  | MVP discovery |  |  | FinnGen replication |  |  |  |  |
| --- | --- | --- | --- | --- | --- | --- | --- | --- | --- | --- | --- | --- | --- |
| CHR | rsID | Nearest gene | Position | A1 | A2 | EAF | OR | P-value | EAF | OR | P-value | One-sided P-value | FDR BH |
| BMI-unadjusted |  |  |  |  |  |  |  |  |  |  |  |  |  |
| 1 | rs17367240 | LINC01787 | 96960057 | A | G | 0.38 | 0.97 | 2.07E-09 | 0.29 | 0.97 | 1.33E-03 | 6.65E-04 | 1.77E-03 |
| 2 | rs58857776 | LINC01122 | 58890684 | T | C | 0.41 | 0.96 | 5.06E-13 | 0.51 | 0.97 | 2.79E-03 | 1.40E-03 | 2.48E-03 |
| 2 | rs77881454 | THADA | 43757293 | T | A | 0.11 | 1.05 | 6.72E-11 | 0.07 | 1.06 | 1.91E-03 | 9.53E-04 | 1.98E-03 |
| 2 | rs540606 | CAMKMT | 45138507 | A | G | 0.39 | 1.03 | 3.71E-10 | 0.36 | 1.02 | 4.12E-02 | 2.06E-02 | 3.00E-02 |
| 2 | rs11691026 | AC018742.1 | 22131865 | A | G | 0.44 | 1.03 | 2.32E-09 | 0.51 | 1.01 | 3.09E-01 | 1.54E-01 | 1.65E-01 |
| 2 | rs181355045 | LINC01876 | 156998417 | A | C | 0.11 | 1.05 | 2.78E-08 | 0.12 | 1.04 | 1.98E-03 | 9.90E-04 | 1.98E-03 |
| 3 | rs869400 | ETV5 | 185826740 | T | G | 0.20 | 0.95 | 2.63E-13 | 0.16 | 0.95 | 1.34E-04 | 6.72E-05 | 2.15E-04 |
| 4 | rs13107325 | SLC39A8 | 103188709 | T | C | 0.07 | 1.09 | 3.58E-16 | 0.01 | 1.04 | 2.93E-01 | 1.46E-01 | 1.65E-01 |
| 4 | rs10938397 | AC108467.1 | 45182527 | G | A | 0.39 | 1.03 | 1.37E-09 | 0.47 | 1.04 | 5.16E-05 | 2.58E-05 | 1.03E-04 |
| 9 | rs5015933 | GAPVD1 | 128137418 | C | T | 0.51 | 0.97 | 2.61E-09 | 0.55 | 0.95 | 1.91E-08 | 9.55E-09 | 7.64E-08 |
| 10 | rs34872471 | TCF7L2 | 114754071 | C | T | 0.30 | 0.97 | 1.93E-09 | 0.20 | 0.98 | 5.86E-02 | 2.93E-02 | 3.61E-02 |
| 10 | rs201575845 | NT5C2 | 104861271 | A | AT | 0.10 | 1.05 | 2.72E-08 | NA | NA | NA | NA | NA |
| 12 | rs2277339 | PRIM1 | 57146069 | G | T | 0.12 | 0.95 | 9.05E-10 | 0.13 | 0.93 | 3.44E-07 | 1.72E-07 | 9.17E-07 |
| 13 | rs201770 | DLEU1 | 51047531 | G | A | 0.22 | 0.97 | 1.32E-08 | 0.30 | 1.00 | 9.55E-01 | 5.23E-01 | 5.23E-01 |
| 15 | rs283789 | SCAPER | 76777577 | G | A | 0.36 | 0.97 | 2.61E-08 | 0.33 | 0.98 | 5.31E-02 | 2.65E-02 | 3.54E-02 |
| 16 | rs1558902 | FTO | 53803574 | A | T | 0.33 | 1.08 | 3.80E-47 | 0.42 | 1.10 | 1.54E-23 | 7.70E-24 | 1.23E-22 |
| 17 | rs58879558 | MAPT | 44095467 | C | T | 0.20 | 1.04 | 8.22E-10 | NA | NA | NA | NA | NA |
| 19 | rs429358 | APOE | 45411941 | C | T | 0.15 | 0.95 | 3.40E-11 | 0.18 | 0.97 | 1.75E-02 | 8.73E-03 | 1.40E-02 |
| BMI-adjusted |  |  |  |  |  |  |  |  |  |  |  |  |  |
| 1 | rs79950770 | TBX15 | 119539797 | T | C | 0.09 | 1.06 | 2.93E-10 | 0.16 | 1.03 | 6.15E-03 | 3.08E-03 | 4.61E-03 |
| 2 | rs72618639 | NRXN1 | 50795733 | G | C | 0.02 | 0.88 | 5.02E-09 | 0.03 | 1.01 | 6.83E-01 | 6.58E-01 | 6.58E-01 |
| 8 | rs34179442 | SNTG1 | 50900444 | A | G | 0.47 | 0.97 | 3.74E-08 | 0.68 | 0.97 | 4.58E-03 | 2.29E-03 | 4.58E-03 |
| 12 | rs2277339 | PRIM1 | 57146069 | G | T | 0.12 | 0.96 | 1.38E-08 | 0.13 | 0.93 | 2.29E-08 | 1.14E-08 | 6.87E-08 |
| 13 | rs592333 | DLEU1 | 51340315 | G | A | 0.46 | 1.03 | 4.68E-08 | 0.52 | 1.04 | 4.48E-05 | 2.24E-05 | 6.72E-05 |
| 15 | rs4886823 | SCAPER | 76965402 | G | A | 0.27 | 0.96 | 1.30E-09 | 0.24 | 0.99 | 3.06E-01 | 1.53E-01 | 1.84E-01 |

CHR: chromosome; Position: genomic coordinate in genome build hg37; A1: effect allele; A2: non-effect allele; OR: estimated odds ratio per one effect allele; P-value: two-sided p-value computed based on the Wald test using a reference normal distribution.
\* MVP sample size: N=568,576 in BMI-unadj analysis; N=559,070 in BMI-adj analysis
\* FinnGen sample size: 27,207 cases, 280,720 controls.
One-sided p-values were computed based on the direction of association observed in the discovery MVP study, so that no association may be replicated if the direction of association does not match across discovery and replication. FDR BH refers to False Discovery Rate adjusted p-values computed using the Benjamini-Hochberg method.

**Figure 3:**
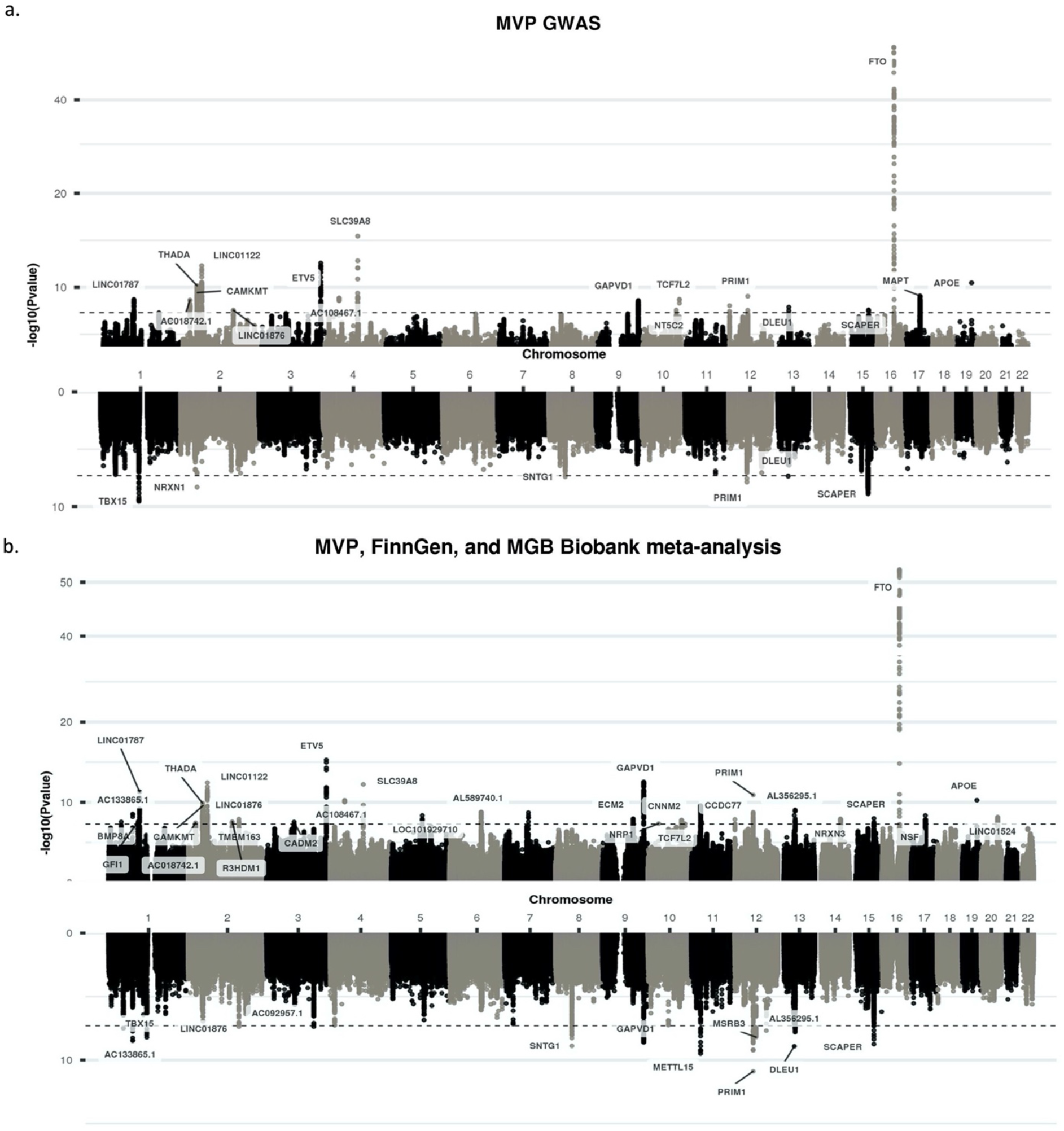
Miami plots of the main MVP analyses and of the MVP, FinnGen, and MGB Biobank meta-analyses. Panel (a) provides a Miami plot from the primary MVP GWAS and panel (b) provides a Miami plot from the meta-analysis of MVP, FinnGen, and MGB Biobank. In each Miami plot, the top part provides results from BMI-unadj analysis, and the bottom from BMI-adj analysis.

Two of the replicated associations are in intergenic regions and associated with long non-coding RNAs, tagged by index SNPs rs17367240 (LINC01787) and rs58857776 (LINC01122). Rs17367240 (replication p-value = 0.001) was not reported in the Type 2 Diabetes Knowledge Portal, while rs58857776 (replication p-value = 0.003) was previously associated with BMI, as well as insomnia and naps (49–51). Rs77881454 (replication p-value 0.002) is in the *THADA* (THADA Armadillo Repeat Containing) gene and was previously associated with glycemic traits (52). It was also associated with p-value=0.004 with minimum oxyhemoglobin saturation during sleep (53). Another replicated SNP at nominal significance (replication p-value=0.04), rs540606, is near the *CAMKMT* (Calmodulin-Lysine N-Methyltransferase) gene, thought to be involved in calcium signaling (54). It was previously reported to be associated with dietary patterns (55). Rs869400 in the *ETV5* (ETS Variant Transcription Factor 5; replication p-value = 1×10-4) was previously reported as associated with BMI, cholesterol, and kidney function (51,56,57). Rs10938397 (replication p-value 5×10^−5^) was also reported as associated with BMI, DM, and HDL (51,56,58). Rs79950770 at the *TBX15* gene (T-box transcription factor 15, encodes transcription factors associated with development; replication p-value=0.006) was associated with OSA in BMI-adj analysis. It was previously associated with waist-to-hip ratio in BMI-adj analysis. Finally, rs34179442 in the *SNTG1* gene (Syntrophin Gamma 1; replication p-value=0.005) was not associated with any trait at the Type 2 Diabetes Knowledge Portal at the genome-wide significance level. Its strongest association was with the OSA phenotype “minimum oxyhemoglobin saturation during sleep” (p-value=8×10^−5^, (26)), with the A allele associated with higher saturation, matching the direction of association of the variant with OSA observed in MVP.

Of the genome-wide significant associations, 12 and 5 of the BMI-unadj and BMI-adj associations had evidence of association with snoring (17), defined as p-value<0.05 and the same direction of associations as with OSA in the MVP GWAS (**Supplementary Table 3**).

### OSA GRS based on BMI-adj analysis is associated with OSA in the MGB biobank in BMI-adj analysis

To further assess replication and generalization of estimated associations, we constructed four genetic risk scores (GRS) in the MGB Biobank, based on the associations reported in **Table 1**: separately based on BMI-adj and BMI-unadj analyses, and for each analysis, one weighted and one unweighted (summation of OSA risk alleles) GRS. MGB Biobank participant characteristics are provided in **Supplementary Table 4**. Complete association analysis results are provided in **Supplementary Table 5**. Weighted GRS (wGRS) had stronger association with OSA compared with the unweighted GRS. The wGRS based on results from BMI adjusted and BMI unadjusted analysis were weakly correlated: Pearson R^2^=0.16, computed over the full MGB analytic sample. **Figure 4** visualizes the estimated associations of the wGRS with OSA in MGB biobank across sex strata and with and without adjusting the analysis in MGB biobank to BMI. Associations tend to be stronger in males compared to females, though not substantially. When adjusting the MGB analysis to BMI, the association of the wGRS that was constructed based on analysis unadjusted to BMI becomes weak and statistically not significant. However, the association of wGRS constructed based on BMI-adj GWAS is generally nominally significant (p-value<0.05) both with and without BMI adjustment and tend to be stronger in BMI-adj association analysis.

**Figure 4:**
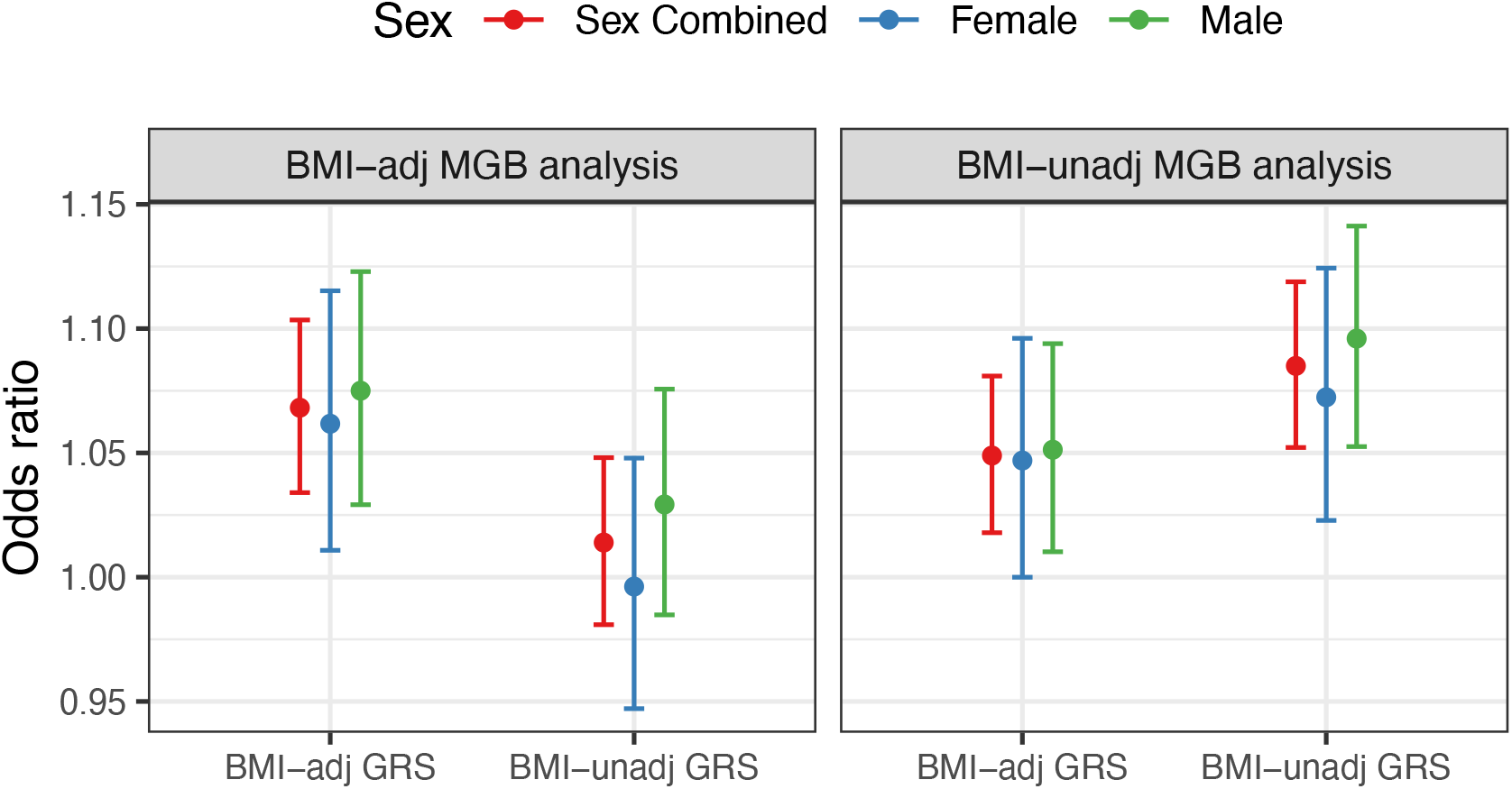
Association of OSA GRSs with OSA in the MGB Biobank. The figure provides estimated odds ratio (OR) per 1 standard deviation (SD) increase in OSA GRSs constructed based on BMI-adjusted GWAS (“BMI_adj”) and based on BMI-unadjusted GWAS (“BMI_unadj”) for OSA in MGB biobank, estimated in BMI-adj (BMI-adjusted analysis; left) and in BMI-unadj (analysis that does not adjust for BMI; right). For GRSs were weighted by the estimated log-OR in MVP.

### Heritability estimates are higher within HARE and biological sex groups suggesting genetic heterogeneity

We performed multiple GWAS in MVP: stratified by sex and HARE group (which were later meta-analyzed for the main results), and in secondary analysis, stratified by obesity. We used the resulting summary statistics to assess evidence of heterogeneity in the genetic basis of OSA across different groups that have known differences in OSA epidemiology.

**Supplementary Figure 4** demonstrates the estimated heritability across sex and HARE groups. Estimated heritability tend to be lower in the multi-population analysis compared to each of the HARE group-specific estimates, and similarly lower in sex-combined analysis compared to sex-specific analyses. Comparing OSA heritability with and without adjustment to BMI, heritability estimates tend to be 5-10% lower when adjusted to BMI. Focusing on the largest HARE groups, heritability is higher in Black male individuals compared White male individuals. Estimated heritability in Hispanic individuals tend to be intermediate between the White and Black estimates. Heritability estimates based on Asian individuals tend to be high, but the level of uncertainty is also very high, due to their low sample size in the analysis.

### Widespread differences in genetic associations with OSA by sex

We compared genetic associations with OSA between males and females. For each variant, we tested the difference in estimated effect size (log OR) between the two groups. The results were striking, and in line with the well-known differences in OSA prevalence between biological sexes through the life course: pervasive sex differences in genetic associations. **Supplementary Figure 5** provides the Manhattan and qq-plots from sex-differences GWAS, demonstrating widespread significant sex differences in genetic associations with OSA. **Figure 5** visualizes the estimated associations (log ORs) of the minor alleles in males and in females across categories of SNPs defined by their p-value in test of sex differences, based on BMI-adj analysis. **Supplementary Figure 6** provides similar results for BMI-unadj analysis. For the variants with most significant associations, all estimated effects in females are protective. This pattern persists as the sex-difference p-value increases, although with progressively fewer variants exhibiting a protective effect in women. We manually annotated the genome-wide significant sex-different SNPs (after clumping, 70 SNPs from BMI-unadj and 68 SNPs from BMI-adj analysis, partly overlapping) using the Type 2 Diabetes Knowledge Portal, reporting associations with p-values<0.001 with any trait (**Supplementary Table 6**). **Supplementary Figure 7** visualizes the number of previously reported SNP associations at this significance level for OSA sex-different SNPs by trait category, based on BMI-adj and BMI-unadj analyses. In BMI-unadj analysis, 7 of the 70 OSA sex-different SNPs were previously associated with glycemic traits, and 4 were associated with cardiovascular traits. For each of the anthropometric, ECG, sleep and circadian, and renal traits, there were 3 associated SNPs. Out of the 68 OSA sex-different SNPs in BMI-adj analysis, 8 and 7 where previously associated with cardiovascular and glycemic traits, respectively, and 3 were associated with renal traits.

**Figure 5:**
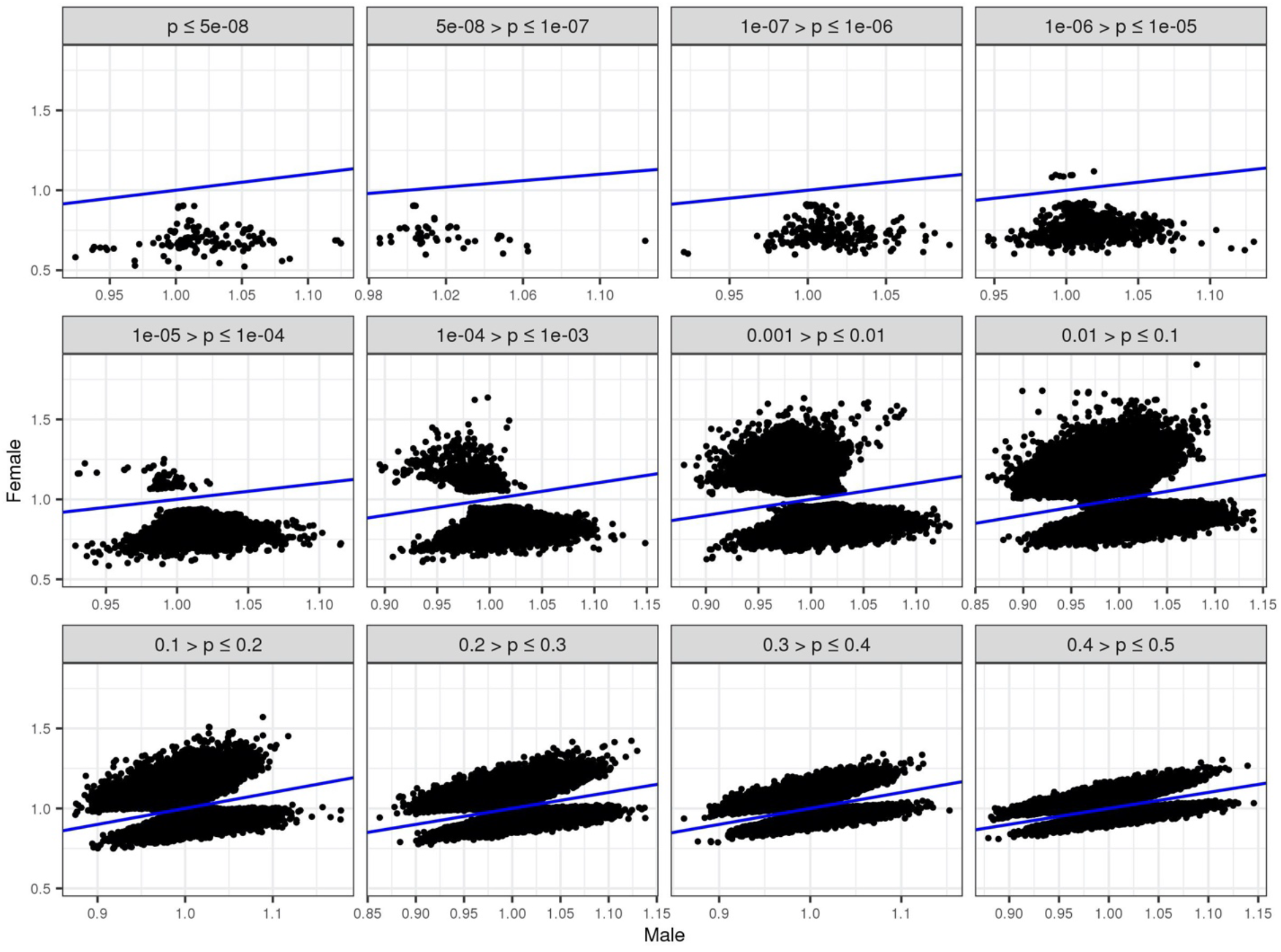
OSA protective genetic associations with OSA in females in SNPs with evident sex interactions. For sets of SNPs defined by their p-value (p) in test of difference in effect sizes between biological sex groups, the figure compares the estimated odds ratio (OR) in males and in females. Depicted results are from BMI-adj analysis. The blue line is the line of identity.

We also performed sex-combined and sex-specific gene-based analyses of the multi-population OSA GWAS. Not surprisingly, due to sample size differences, most associations were driven by the results in males. In BMI-unadj analysis (**Supplementary Table 7)**, the strongest association was of the Fat Mass and Obesity Associated gene (*FTO*), with evidence of association in both males (p-value=3×10^−45^) and females (p-value=1.8×10^−4^). In males, the second strongest association was of the *ETV5* (ETS Variant Transcription Factor 5) gene, which had no evidence of association in females (p-value=0.23). This gene is involved in cellular response to oxidative stress (59) and is mostly expressed in the pituitary gland according to the gene-tissue expression (GTEx) portal (60). The top association in females and only association with FDR p-value <0.05 was with the *SPIDR*, Scaffold Protein Involved in DNA Repair, gene (FDR=0.04; p-value=2.3×10^−6^) and was not associated with OSA in males (p-value=0.08). In GTEx, *SPIDR* is mostly expressed in ovary tissue and in the cerebellar hemisphere. Similarly, in BMI-adj analysis (**Supplementary Table 8**) some of the top associated genes had evidence of association in males only, including, the *SCAPER* and *ETFA* genes (female p-values>0.8). None of the genes passed FDR correction for females in BMI-adj analysis. *SCAPER* (S-Phase Cyclin A Associated Protein In The ER), is highly expressed in testis, tibial artery, and cerebellar hemisphere. *ETFA* (electron transfer flavoprotein subunit alpha) encodes a subunit of the enzyme flavoprotein, which is active in the mitochondria in fatty acid beta-oxidation (61).

We also compared SNP associations between the White and Black HARE groups, as the two largest groups, and tested the difference in estimated effects across groups. Manhattan and qq-plots are reported in **Supplementary Figure 8**, and top results are provided in **Supplementary Table 9**. Compared with the sex-differences GWAS, there are fewer genome-wide significant associations. After clumping of SNPs, there were 20 genome-wide significant associations (8 in BMI-adj and 12 in BMI-unadj). In annotation of these SNPs in the Type 2 Diabetes Knowledge Portal, most of these SNPs were not previously reported (p-value>0.001 in all reported GWAS), while 7 of them were previously associated (p-value<0.001) with a range of phenotypes, including glycemic and anthropometric traits, and snoring.

### No evidence for heterogeneity in top associations when accounting for statistical power

To estimate the evidence of generalization of associations between groups, for each HARE, sex, and obesity group, we identified genome-wide significant associated SNPs, and evaluated the evidence of OSA association of these SNPs within other groups. **Supplementary Tables 10** and **11** report genome-wide significant SNP associations based on each of the main and secondary analyses, separated by BMI-adj and BMI-unadj analyses, and further reports their associations across all strata-specific analyses. The results from analysis of generalization across groups are provided in **Supplementary Figure 9**, where we provide the number of observed associations in each stratum when using the genome-wide significance threshold, and when using the nominal 0.05 threshold. The figure also annotates the expected number of observed associations at the 0.05 threshold when accounting for a stratum’s sample size, number of cases, and allele frequencies. While some associations are not observed in some groups, for example, none of the 3 White HARE group-specific associations were observed in the Black HARE group, the number of generalizations at the 0.05 p-value level always exceeded the expected number of generalizations computed using power analysis.

### Genetic correlations of OSA with cardiometabolic phenotypes and evidence of pleiotropy with and without accounting for BMI

We estimated the genetic correlation between OSA and a range of phenotypes, including sleep, lipids, blood pressure, diabetes mellitus (DM), and Alzheimer’s disease. For each of the traits we performed both BMI-adj and BMI-unadj GWAS and used the summary statistics to estimate genetic correlations between OSA and each of the traits with and without accounting for BMI. The results are visualized in **Figure 6**. The estimated genetic correlations between OSA and blood pressure, lipids, and DM reduced substantially once accounting for BMI and often lost statistical significance. Still, the estimated genetic correlation between OSA and DM in BMI-adj analysis 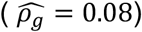 was statistically significant (p-value=0.03). There were multiple statistically significant genetic correlations between OSA and sleep traits, and these often substantially changed between BMI-adj and BMI-unadj analysis. For example, between OSA and insomnia in BMI-unadj we estimated 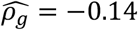 (p-value=0.02) while in BMI-adj 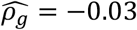 (p-value=0.66); between OSA and “trouble falling asleep” in BMI-unadj we had 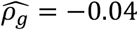 (p-value=0.29) while in BMI-adj 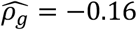 (p-value=5×10^−4^); and between OSA and “excessively sleepy” in BMI-unadj 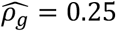 (p-value=3.7×10^−7^) while in BMI-adj 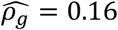 (p-value=0.01).

**Figure 6:**
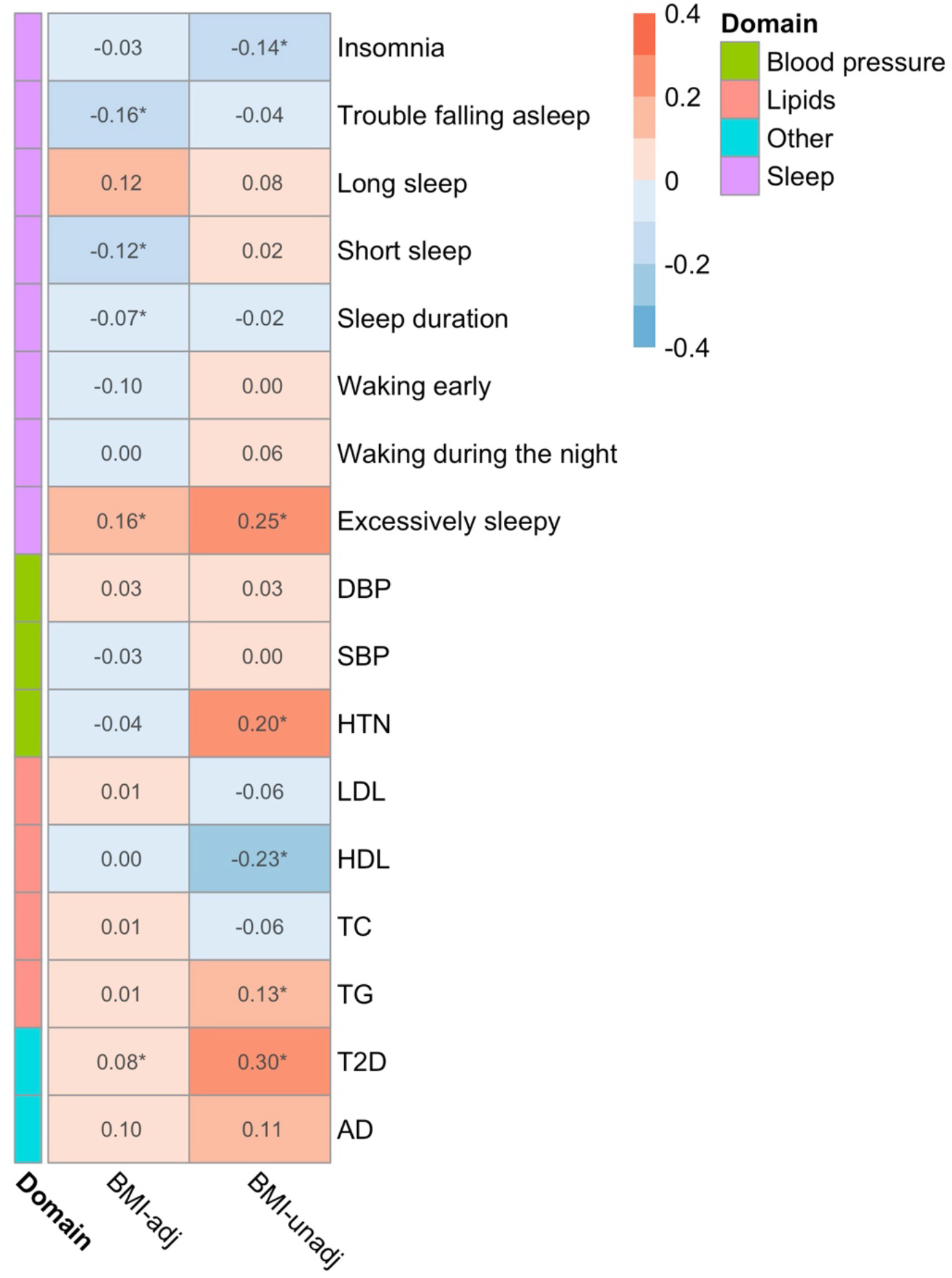
Genetic correlations of OSA with cardiometabolic traits are driven by BMI. Estimated genetic correlations between OSA and other phenotypes, in BMI-adj and BMI-unadj analyses. For each pair of phenotypes, genetic correlations were compute using LDSC based on summary statistics from GWAS, restricted to HapMap SNPs. Insomnia was inferred from the EHR, while other sleep measures were self-reported from the baseline MVP survey. DBP: diastolic blood pressure; SBP: systolic blood pressure; HTN: hypertension; LDL: low-density lipoprotein cholesterol; HDL: high-density lipoprotein cholesterol; TC: total cholesterol; TG: triglycerides; T2D: type 2 diabetes; AD: Alzheimer’s disease. * indicates p<0.05.

We also considered the evidence of pleiotropy in genome-wide significant OSA SNPs (**Supplementary Tables 12** and **13**). At the genome-wide significance level, in BMI-unadj analysis many OSA-SNP associations were identified with other traits, including insomnia/sleepiness, blood pressure, coronary artery disease, lipids, glycemic traits, and as well as alcohol use and smoking. When considering BMI-adj analyses, fewer (six) SNPs are considered. None of these SNPs is associated with the considered outcomes at the 5×10^−8^ level in BMI-adj analysis. Rs2277339 near the *PRIM1* gene is associated at the nominal level (p-value<0.05) with multiple cardiovascular, lipids, and glycemic traits, as well as alcohol, smoking, and depression. *PRIM1* gene is the DNA Primase Subunit 1.

### Secondary GWAS: meta-analysis of MVP and other biobank-based GWAS

We meta-analyzed the primary GWAS from MVP, the publicly available FinnGen Freeze 7 GWAS summary statistics, and a GWAS of OSA from the MGB Biobank. **Figure 3 panel b** provides the Miami plot, and **Supplementary Table 14** provides the top (clumped, genome-wide significant) associations. There were 31 and 12 genome-wide significant associations with OSA in BMI-unadj and BMI-adj analyses, respectively. **Supplementary Tables 15** and **16** provide replication and proxy-replication look ups of the genome-wide significant SNPs in related GWAS: snoring (BMI-adj and BMI-unadj) from the UKBB (17), apnea-hypopnea index (9), minimum and average oxyhemoglobin saturation during sleep (26), both studies are meta-analyses of multiple cohorts, largely overlapping, with dedicated sleep exams, OSA from the PAN-UKBB GWAS summary statistics database (https://pan.ukbb.broadinstitute.org/), and OSA from a GWAS in a Han Chinese population reported by Xu et al. (11). Supplementary Table 17 summarizes the overlap between this GWAS, the MVP-only GWAS, the previously published FinnGen GWAS (16), and the OSA GWAS using multi-trait analysis with snoring (18), and the evidence of replication and proxy-replication. This analysis detected 15 associations regions that were not previously reported in the other GWAS. Of these, 5 regions had evidence of association in the Pan UKBB sleep apnea GWAS, and 5 additional regions had evidence of association with snoring.

## Discussion

We performed GWAS of OSA in a large dataset of N=568,576 MVP participants, and further performed a meta-analysis of MVP, FinnGen, and the MGB Biobank for a total of 916,696 individuals. We used the MVP dataset to comprehensively study the known race/ethnic- and sex heterogeneity of OSA from a genetic perspective, and to glean insight into the role of BMI in the association of OSA with other phenotypes. We see evidence that many OSA genetic loci are associated with other sleep phenotypes, affirming the contribution of OSA to impaired sleep. We also identified and replicated or proxy-replicated (using other sleep phenotypes) multiple loci that were not yet reported for sleep phenotypes. We show that a GRS based on BMI-adj OSA GWAS is associated with OSA even when adjusting for BMI in an independent dataset, confirming a role of genetics in OSA etiology independent of BMI. Finally, we see pervasive sex differences in genetic associations with OSA, with most significant differences associated with lower OSA risk in women than in men, suggesting that the strong sex differences in OSA prevalence and severity may be related in part to sex differences in the phenotypic manifestation of these genetic variants.

Almost all OSA variant associations have some level of associations in a GWAS of snoring from the UK Biobank (17). While this is expected, as snoring is a cardinal symptom of OSA, there were also a few SNP associations that were not associated with snoring in the UKB GWAS. For example, rs1373285, near a long non-coding RNA, was discovered in the MVP, FinnGen, and MGB Biobank meta-analysis in BMI-adj analysis (OR of the A allele = 1.03, p-value=1.6×10^−8^) and the association was replicated in the Pan-UKBB GWAS of sleep apnea (OR of the A allele = 1.08, p-value=3.3×10^−4^). The A allele of this SNP was previously associated with higher odds or frequent insomnia symptoms in a UKB GWAS (50). This suggests that OSA may lead to insomnia, likely due to poor sleep quality, in some individuals who may not snore. Another SNP, rs4953923 was associated with OSA in BMI-unadj analysis (OR of the T allele = 0.96, p-value=3.1×10^−8^) and did not proxy-replicate for snoring. It demonstrated weak evidence of replication in the Pan-UKBB sleep apnea GWAS (OR of the T allele = 0.94, p-value=0.04). Its strongest association with a sleep phenotype, as seen in the Type 2 Diabetes Knowledge Portal, was with chronotype: T allele was associated with lower likelihood of being a morning person. Rs13107325 (Near the *SLC39A8*, solute carrier family 39 member 8 gene) was discovered and replicated in the Pan-UKBB OSA GWAS, with its T allele increasing risk of OSA. This SNP as well was not associated with snoring (p-value=0.36) but it was associated with both short sleep duration (2.5×10^−13^) and continuous sleep duration (62), with the T allele associated with shorter sleep. The protein encoded by this gene is found in the mitochondria, and has a role in the cellular import of zinc at the onset of inflammation (63). Within MVP, we computed the genetic correlation between OSA and other sleep phenotypes available via participant survey, in BMI-adj and BMI-unadj analyses. For most self-reported sleep phenotypes, estimated genetic correlations substantially changed between the two analyses. For example, the genetic correlation of OSA with short sleep was 0.02 in BMI-unadj and -0.12 in BMI-adj analyses. Notably, results from this analysis may not exactly mirror patterns observed in an association of any single variant, and therefore while we observe that some variants both increase risk of OSA and of insomnia, e.g., it is possible that an overall genetic correlation, averaging genetic contributions over genetic variants from across the genome, may be negative. More work is needed to identify commonalities and differences in genetic underpinnings of sleep traits.

Gender and sex differences are well known for OSA. Previous work reported that the association of OSA with body fat distribution measures differ by sex (64), and gender differences in OSA phenotypes, such as respiratory event length and apnea hypopnea index during non-REM sleep (65,66). This may be on top of well-established gender and sex differences in anthropometric measures (67), and in the genetics underlying such measures, e.g., BMI, waist-to-hip ratio, neck circumferences, and others (68,69). Mechanisms leading to gender and sex differences in genetic associations may include amplification of gene expression due to hormone levels, contributions of the X-chromosome, differential genetic liability distributions, and gene-environment interaction, where exposures that tend to have differential distribution across genders modify genetic effects (70,71). In this work we see genetic differences across sex groups throughout the autosomes. Annotation of the genome-wide significant sex-different OSA variants based on previously-published associations did not reveal specific patterns, though we saw many glycemic trait and cardiovascular-related associations. This observation may be related to reported sex differences in diabetes sensitivity (72). Notably, in previous work we saw that glycemic traits have a causal effect on OSA, as suggested by Mendelian randomization analysis (73). Thus, gender and sex differences in glycemic traits may be related to the observed sex differences in OSA genetics. Future work should further dissect gender and sex differences in OSA and its determinants, and study the role of sex chromosomes in sex differences in OSA.

We studied the genetic evidence for the heterogeneity of OSA across HARE groups defined by race/ethnicity and genetics, as well as by obesity. First, heritability estimates within strata tend to be higher compared to estimates computed over groups that combine multiple strata (multi-population, as well as combined sex groups). A similar phenomena was previously reported (74) in a study of sex differences: the authors reported that estimated heritability in sex-combined analysis tended to be lower compared to estimated heritability than that estimated within sex groups, and this pattern became more pronounced when the genetic correlations between male and female genetic effects became lower. Thus, the heritability analysis supports the existence of some differences in OSA genetic architecture between groups. Yet, in an analysis weighing the evidence for generalization of genome-wide significant associations across strata, when considering the power to detect associations, we could not observe any difference between groups. In contrast, rates of generalizations at the p-value<0.05 were sometimes higher than expected according to the power analysis. Nevertheless, this analysis is limited because the majority of MVP participants (67%) are White males, so that power to discover associations that are specific to other groups is lower. Our analysis of differences between White and Black HARE groups OSA associations did identify a handful of associations, though to much lesser degree compared with sex-different OSA SNPs. Critically, these differences can be due to different exposures modifying genetic effects, i.e., gene by environment interactions. In fact, two of the SNP associations that had HARE differences were within OSA-associated genes: *FTO* and *PRIM1*, suggesting that there may be differences in effect size of known genes. This could also be due to gene-gene interactions (and not only gene-environment interactions).

Strengths of this study include large sample size, high prevalence of OSA that is close to the population level rather than the lower level often observed in biobanks, where lower levels are due to underdiagnosis and thus cases are often misclassified as controls. Also, we performed BMI-adj analysis, improving upon the largest previously-published GWAS of OSA from FinnGen (6) that did not provide genome-wide results from BMI-adj analysis. We further studied the genetic correlation between OSA and cardiometabolic and other phenotypes with and without accounting for BMI. Another strength is the investigation of sex differences. Study limitations include the population not being representative: participants are U.S. Veterans and predominately males. Phenotypes are based on medical records, and as such are prone to various selection biases. For example, some individuals may be treated in other healthcare systems so that covariate and treatment information may not be complete. Individuals are possibly underdiagnosed for OSA as well as other comorbidities, due to self- or doctor-selection for diagnosis and treatment. However, importantly, the VA healthcare system provides free treatment for OSA if needed, therefore the likely proportion of patients with undiagnosed OSA among controls is expected to be lower compared to other biobanks.

In summary, we report a large GWAS of OSA in MVP, assessing in detail the genetic heterogeneity of OSA across sex and HARE groups, and its relation to other phenotypes, and a second GWAS meta-analysis of MVP, FinnGen, and the MGB Biobank, leveraging large available sample sizes for increased discovery power. Future work should build upon these findings to develop strategies to identify undiagnosed individuals who likely have OSA and may benefit from treatment, potentially in a sex-specific manner.

## Data Availability

Full GWAS summary statistics are being deposited on dbGaP (https://www.ncbi.nlm.nih.gov/gap/) under the MVP accession (phs001672). FinnGen GWAS summary statistics (version 7) are available from the FinnGen portal (https://www.finngen.fi/en/public-release-finngen-data-freeze-7-results-and-summary-statistics). MGB Biobank data are available to MGB investigators. Summary statistics for sleep-related phenotypes were access through the Type 2 Diabetes Knowledge Portal (https://t2d.hugeamp.org/). Summary statistics of sleep apnea GWAS from UKBB were accessed via the Broad Institute webpage (https://pan.ukbb.broadinstitute.org/). 

https://www.finngen.fi/en/public-release-finngen-data-freeze-7-results-and-summary-statistics

https://t2d.hugeamp.org/

https://pan.ukbb.broadinstitute.org/

## Ethics statement

MVP received ethical/study protocol approval from the VA Central Institutional Review Board, and written informed consent was obtained for all participants.

## Acknowledgements

We are grateful to the Million Veteran Program participants and staff. This research is based on data from the Million Veteran Program, Office of Research and Development, Veterans Health Administration, and was supported by award #BX004821. This publication does not represent the views of the Department of Veteran Affairs or the United States Government.

## Authors and contributors

Study conceptualisation: T Sofer, DJ Gottlieb. Data curation: M Murray, Y-L Ho, J Huffman, K Cho, PWF Wilson. Formal analysis: N Kurniansyah, M Murray. Funding acquisition: K Cho, PFW Wilson. Supervision: T Sofer. Visualisation: N Kurnianyah. Writing – original draft: T Sofer, N Kurniansyah. Writing – review & editing: M Murray, Y-L Ho, J Huffman, K Cho, PFW Wilson, DJ Gottlieb. T Sofer and N Kurniansyah have directly accessed and verified the underlying data reported in the manuscript. All authors had full access to all the data in the study and accept responsibility to submit for publication.

