## Supplemental text and figures for "Genome-wide association study of obstructive sleep apnea in the Million Veteran Program uncovers genetic heterogeneity by sex"

#### Supplementary Materials

#### Supplemental Notes

##### Supplemental Note 1: The Mass General Brigham Biobank

Samples, genomic data, and health information were obtained from the Mass General Brigham (MGB) Biobank, a biorepository of consented patient samples at Mass General Brigham.

**DNA samples**

DNA samples are processed from whole blood that was collected as a dedicated research draw or as a clinical discard. Dedicated research samples are aimed to be processed within four hours of collection. Clinical discards are processed 24+ hours after collection. Whole blood is spun to buffy coat with a centrifuge and the buffy coat is stored in a freezer up to several months. The buffy coat is then extracted to DNA. The DNA is then placed in an ultralow freezer (-80oC). Each DNA aliquot contains a minimum of 2 ug of DNA. The concentration varies.

**Genotyping**

Samples have been genotyped using three versions of the biobank SNP array offered by Illumina that is designed to capture the diversity of genetic backgrounds across the globe. The first batch of data was generated on the Multi-Ethnic Genotyping Array (MEGA) array, the first release of this SNP array. The second, third, and fourth batches were generated on the Expanded Multi-Ethnic Genotyping Array (MEGA Ex) array. All remaining data were generated on the Multi-Ethnic Global (MEG) BeadChip.

**Imputation**

Prior to performing imputation, files were converted to VCF format, separated by chromosomes. When multiple probes measured the same genotypes, they were checked for concordance and were set to a missing value if the genotypes did not match. Files were uploaded to the Michigan Imputation Server, and Genotypes were imputed using TOPMed reference panel. Genomic coordinates are provided in GRCh38.

**Quality control**

We performed quality control using PLINK (v2.0. We filtered SNPs with low-quality imputation (r < 0.8), with missing call rates > 0.1, HWE p-value less than $1x10^{-6}$ and MAF <1%.

We computed principal component (PC) using PLINK: we pruned the genotype data using a window size of 1000 variants, sliding across the genome with a step size of 250 variants at a time, filtering out any SNPs with LD $R^2$>0.1. We used unrelated individuals (3rd degree, identified using PLINK) to compute the loadings for the first 10 PCs.

**Phenotypes**

OSA status was extract from the field "Obstructive Sleep Apnea". Age was the current age of participants, and BMI was taken as the median BMI in the health records.

**MGB Genome-wide association study**

We perform association analyses using PLINK v2.00a3LM (xx) in European. Analyses were adjusted for age at the latest OSA ICD code, sex , genetics batch effect, and the first 10 PCs of genetic data. In addition, a BMI adjusted model further adjusted for BMI, using both linear and squared terms. BMI measures were and BMI was taken as the median BMI in the health records.

#### Supplemental Note 2: Non-OSA phenotype definitions in MVP

We used various phenotypes in secondary analyses: genetic correlation analysis, and analysis of OSA-SNP associations with potentially pleiotropic traits to OSA. For each participant, we defined the "index date". For OSA cases, the index data is the date of their first relevant diagnosis code. For OSA controls, it is the date of their last visit.

**Sleep phenotypes**

Insomnia status was inferred using a MAP algorithm applied on relevant ICD codes and elements extracted using natural language processing. All other sleep phenotypes were self-reported based on the baseline questionnaire administered to program participants. Specifically, sleep duration was defined based on the response to the question about hours of sleep in a typical day, with responses ranging from '5 or less' to '10 or more', with increments of 0.5. Long sleep was defined as sleep duration >9, short sleep as sleep duration <6. Trouble falling asleep, waking early, waking during the night, and excessively sleepy items referred to self-reported sleep problems occurring at least half the days of the past year, with responses being yes or no.

**Disease phenotypes**

Alzheimer's disease: a positive case as having at least one inpatient or two outpatient relevant ICD codes based on the index date. Controls were non-cases. Type 2 diabetes: a positive case as having at least one inpatient or two outpatient relevant ICD codes based on the index date, and additionally having at least one prescription for a diabetes medication. Controls were non-cases. Hypertension was computed based on systolic and diastolic blood pressure (SBP, DBP) as described below.

**Clinical event outcomes**

Acute ischemic stroke (referred here as stroke, for short) phenotype was developed as a prediction based on relevant ICD codes from an algorithm trained over 268 chart-revied samples (1). Heart failure with preserved ejection fraction (HFpEF) cases were defined by having at least one inpatient or two outpatient relevant ICD codes by the index date. Controls were non-cases. Coronary heart disease was defined by the MVP data core using a MAP phenotyping algorithm (2).

**Continuous blood pressure phenotypes and hypertension**

Diastolic and systolic blood pressure (DBP, SBP) were extracted from the available date nearest to the index date. Current use of antihypertensive medication was determined based on prescription of medications within the 6 months window prior to the index date. For medication users, SBP and DBP values were raised by 15 and 10mmHg, respectively. Hypertension was defined as SBP$\geq 130$, DBP$\geq 80$, or current use of antihypertensive medications.

**Lab measures: lipids, HbA1c**

These values were taken from lab measurements closest to the index date.

**Smoking phenotypes**

Smoking phenotypes were derived using an algorithm developed by the MVP Data Core. The algorithm yields the probability of being a never, former, or current smoker, and each individual is classified according to the category with the highest probability. The algorithm was developed using a LASSO regression (3) with 10-fold cross validation on various potential predictors of smoking status and using the MVP baseline survey.

**Mental health**

Alcohol use disorder, post-traumatic stress disorder, opioid use disorder, depression, were based on the index date for each individual, with cases defined by having at least one inpatient or two outpatient relevant ICD codes by the index date. Controls were non-cases.

### Supplemental figures

#### Supplementary Figure 1: QQ plots for the OSA GWASs.

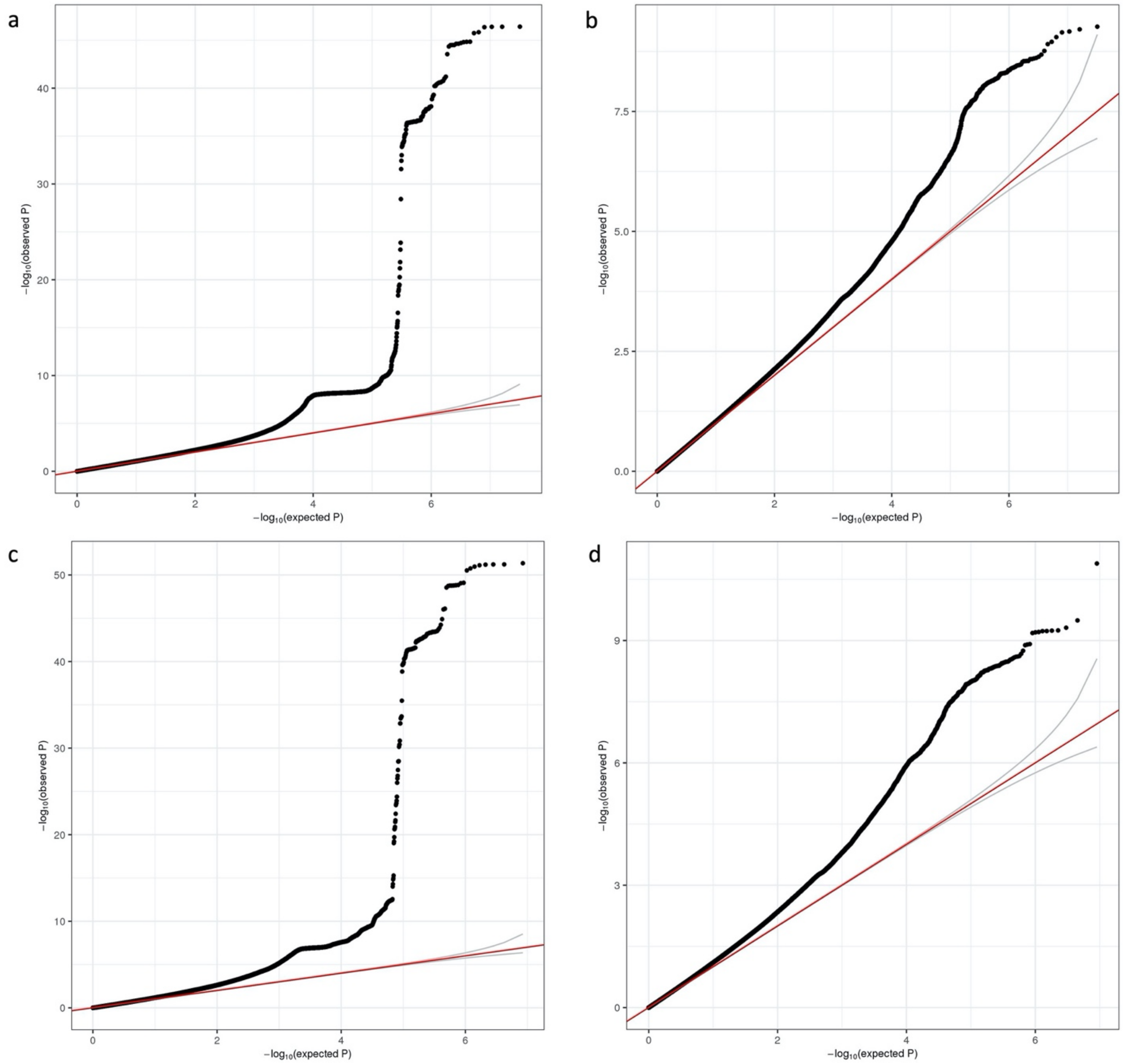

Panel a: qq-plot from the MVP multi-population BMI-unadj GWAS. Panel b: qq-plot from the MVP multi-population BMI-adj GWAS. Panel c: qq-plot from the meta-analysis of MVP, FinnGen, and MGB Biobank BMI-unadj GWAS. Panel d: qq-plot from the meta-analysis of MVP, FinnGen, and MGB Biobank BMI-adj GWAS.

Supplementary Figure 2. Regional association plots from main BMI-unadj analysis

rs17367240 (chr1:96960057:A:G)

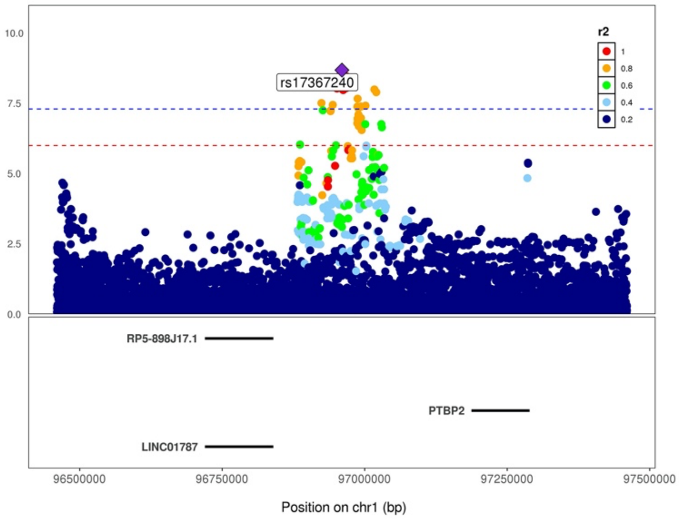

rs58857776 (chr2:58890684:T:C)

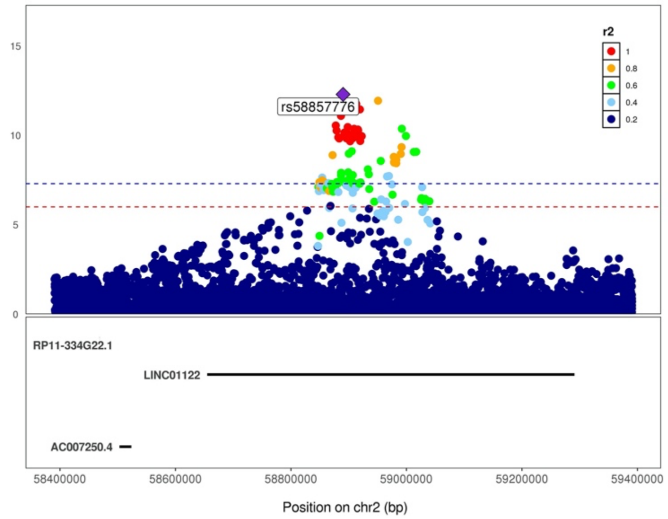

rs77881454 (chr2:43757293:T:A)

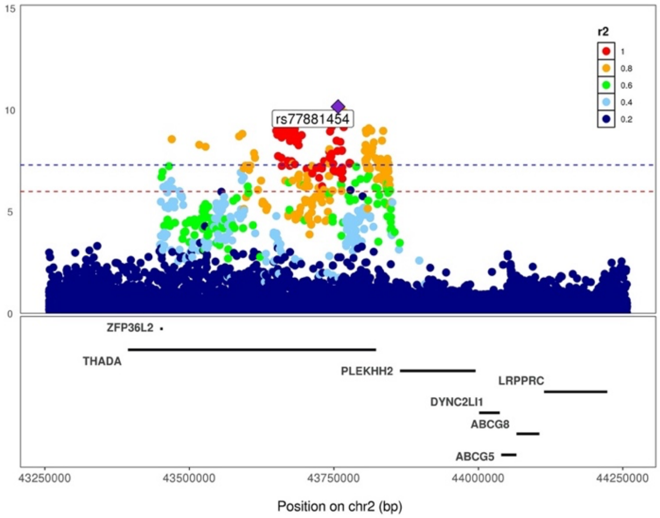

rs540606 (chr2: 45138507:A:G)

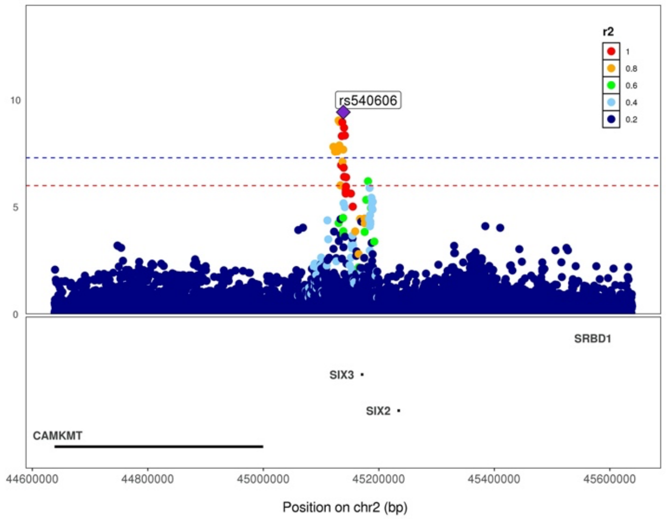

rs11691026 (chr2:22131865:A:G)

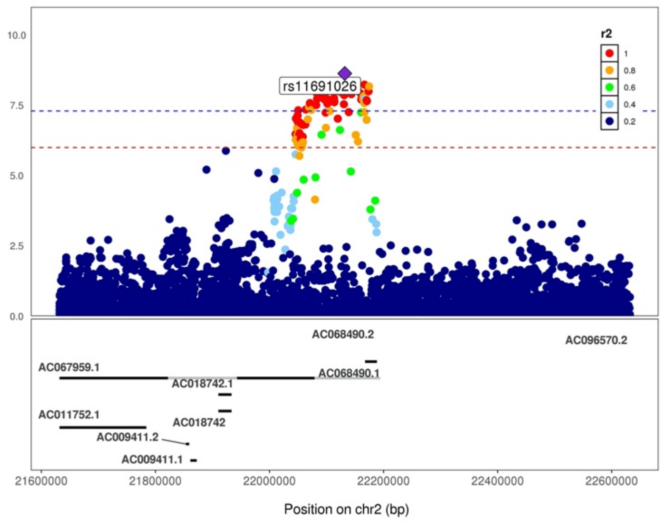

rs181355045 (chr2:156998417:A:C)

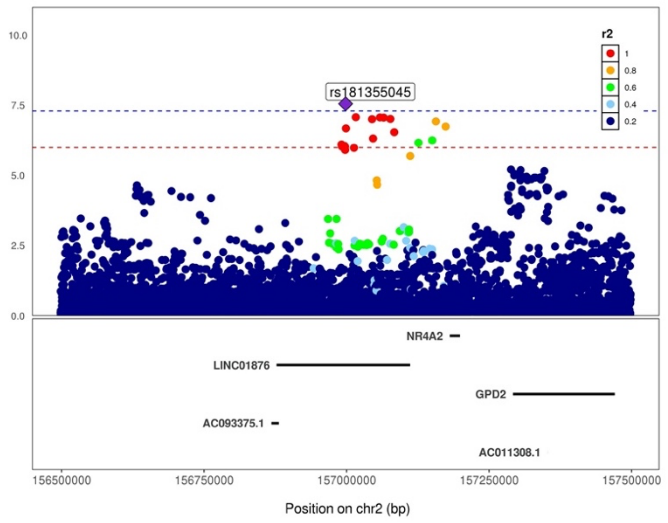

rs869400 (chr3:185826740:T:G)

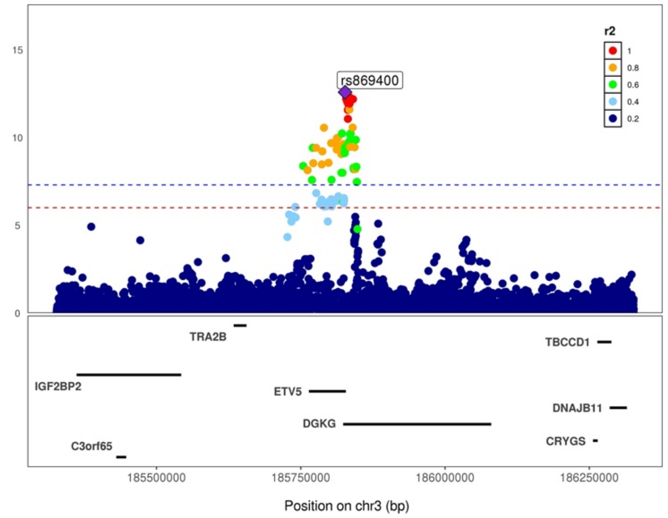

rs13107325 (chr4:103188709:T:C)

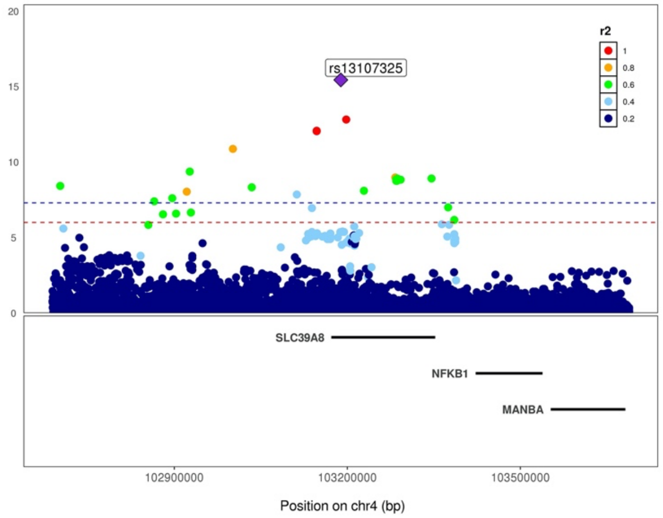

rs10938397 (chr4:45182527:G:A)

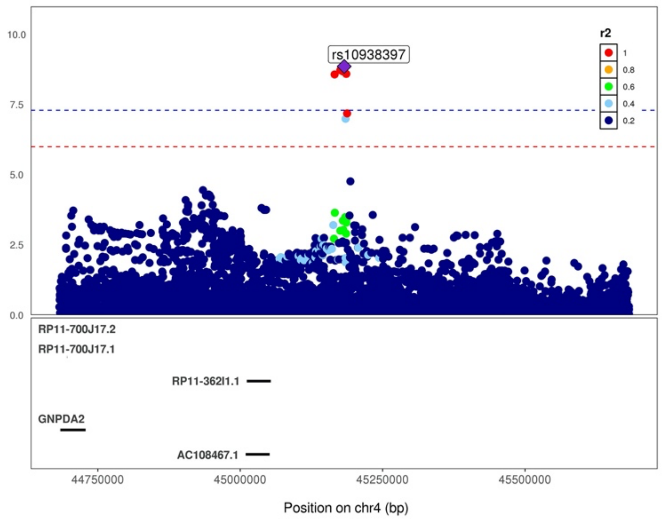

rs5015933 (chr9:128137418:C:T)

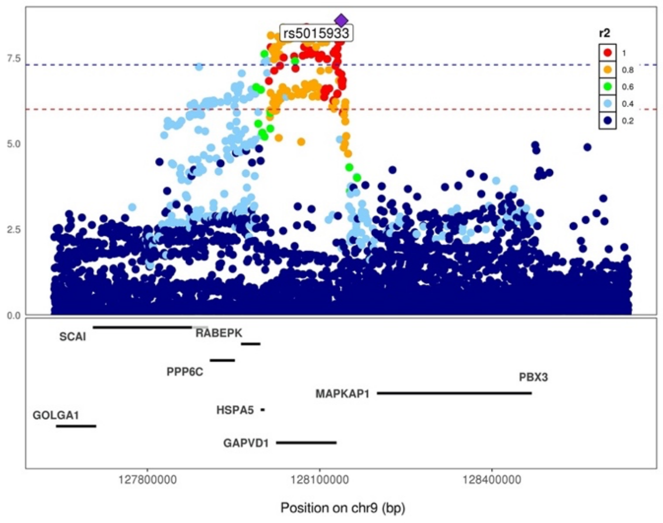

rs34872471 (chr10:114754071:C:T)

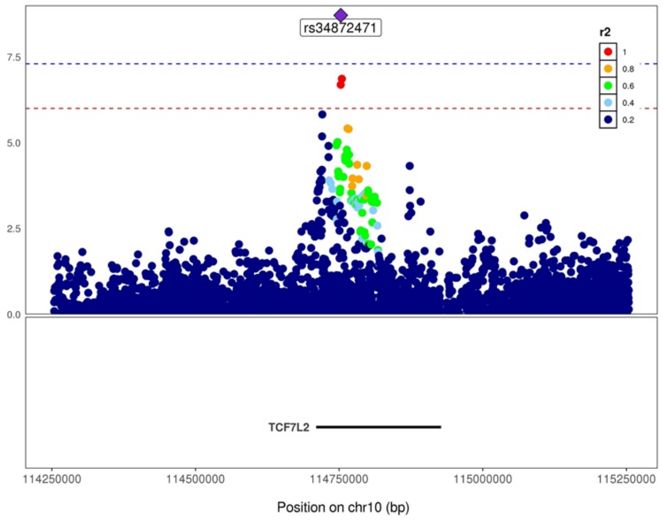

rs201575845(chr10:104861271:A:AT)

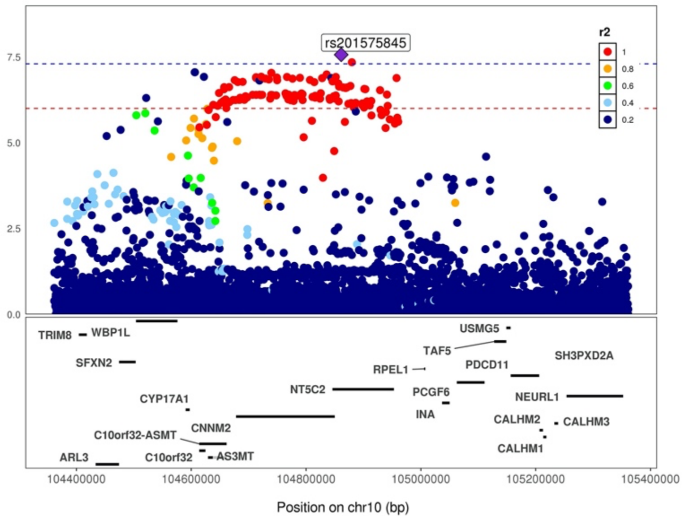

rs2277339 (chr12:57146069:G:T)

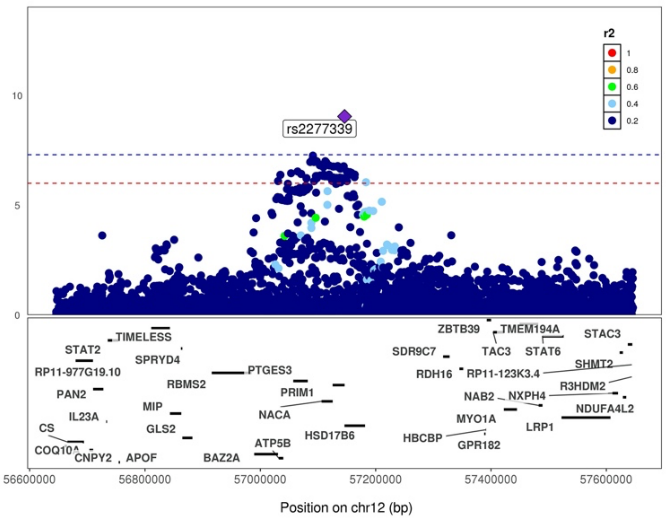

rs201770 (chr13:51047531:G:A)

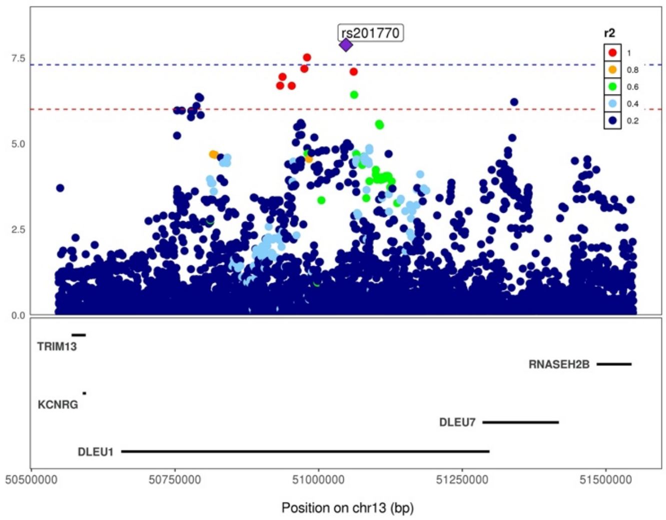

rs283789 (chr15:76777577:G:A)

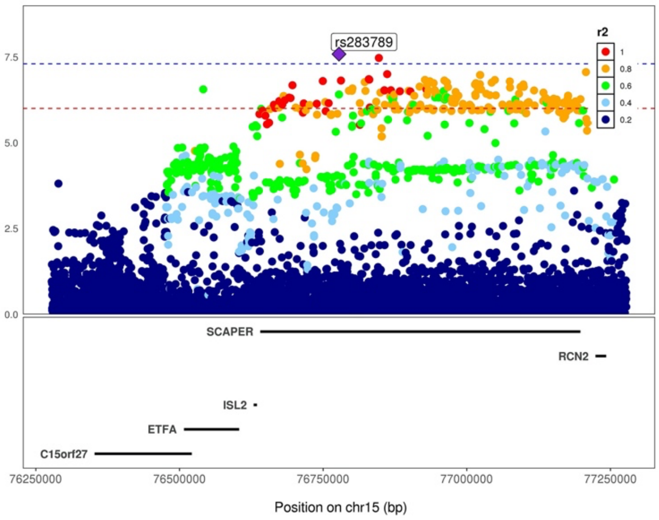

rs1558902 (chr16:53803574:A:T)

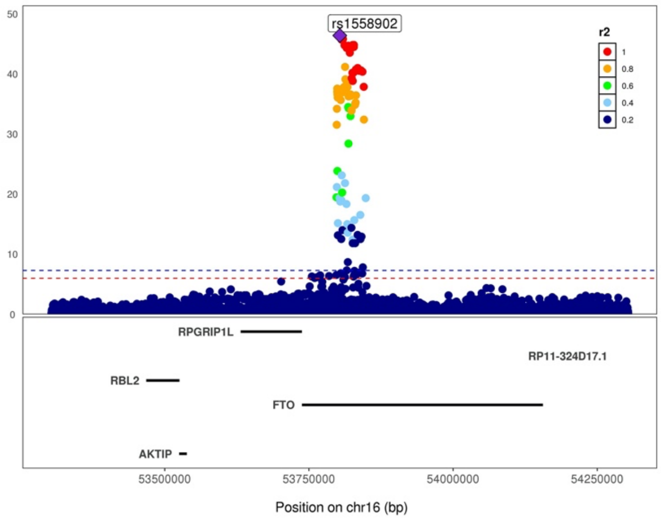

rs58879558 (chr17:44095467:C:T)

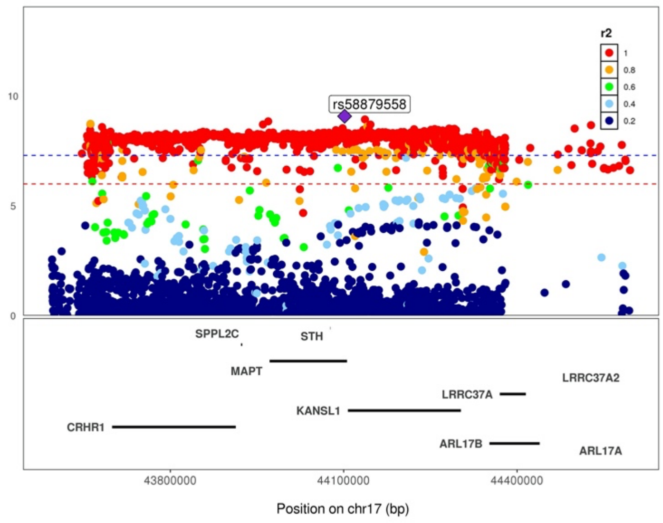

rs429358 (chr19:45411941:C:T)

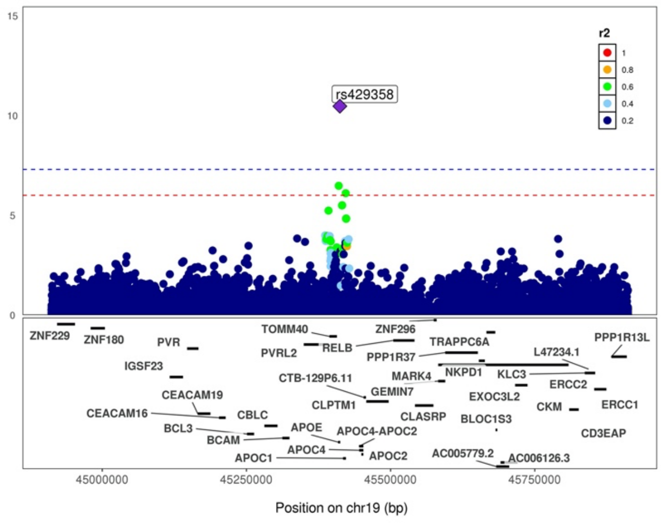

#### Supplementary Figure 3. Regional association plots from main BMI-adj analysis

rs79950770 (chr1:119539797:T:C)

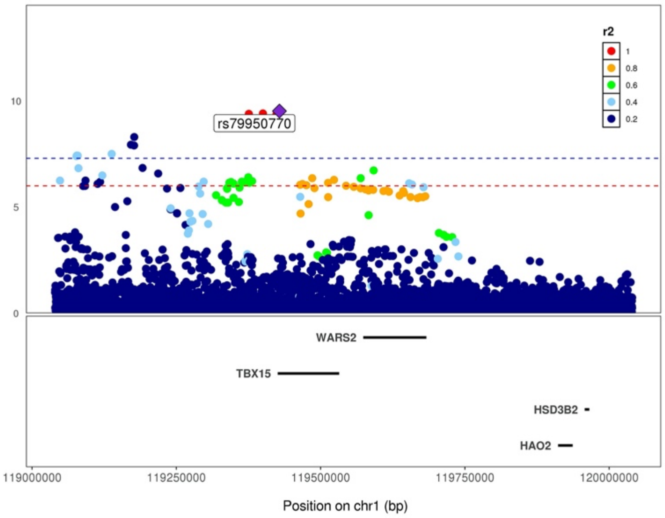

rs72618639 (chr2:50795733:G:C)

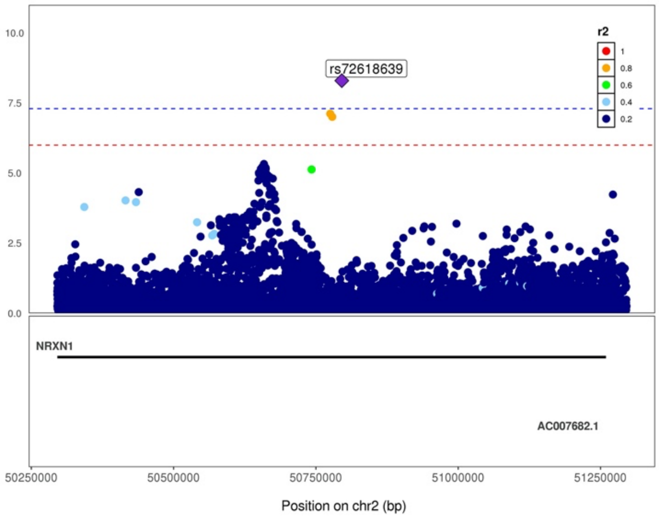

rs34179442 (chr8: 50900444:A:G)

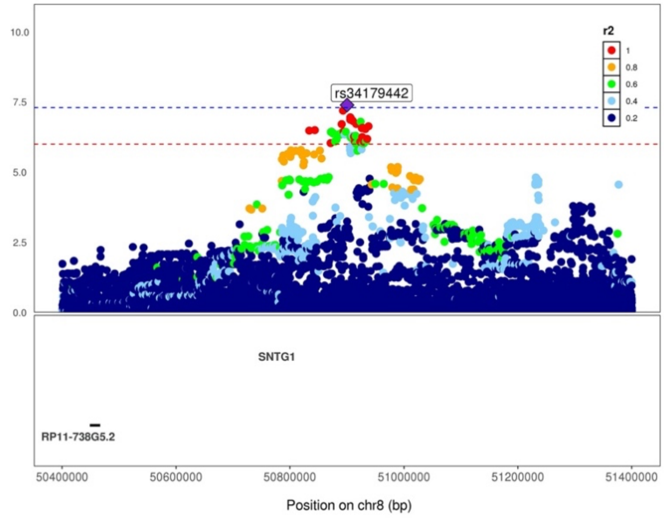

rs2277339 (chr12:57146069:G:T)

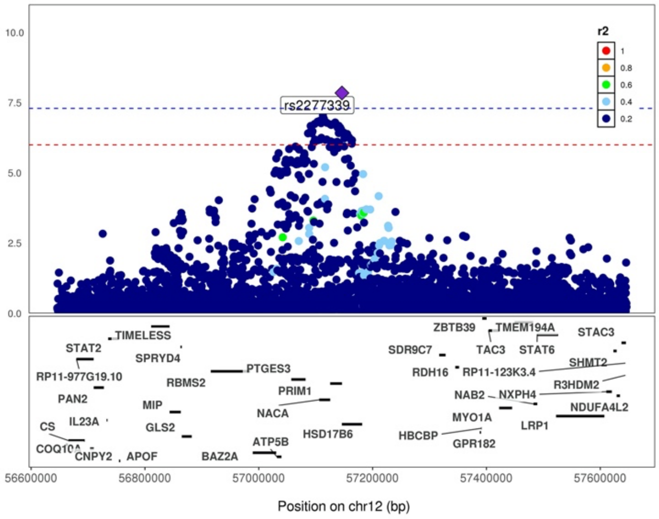

rs592333 (chr13:51340315:G:A)

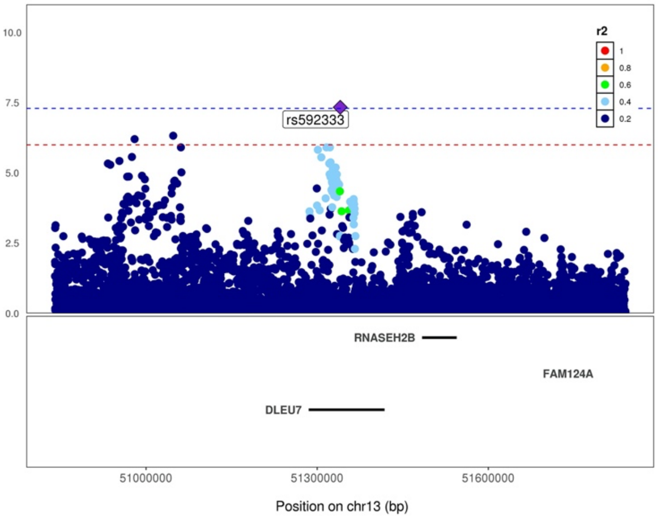

rs4886823 (chr15:76965402:G:A)

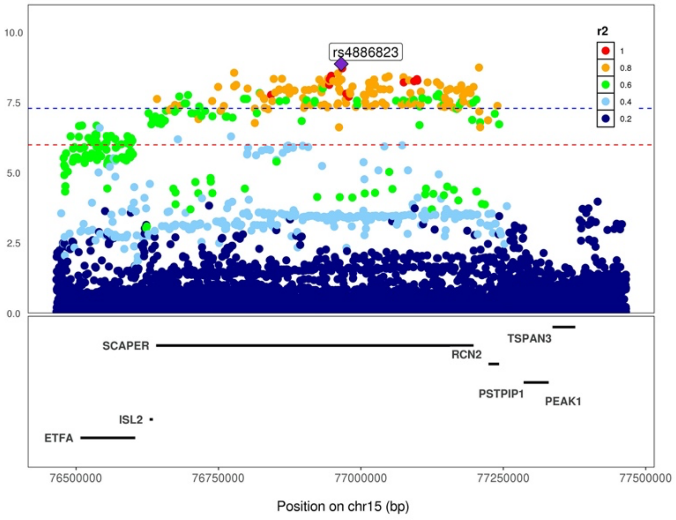

Supplementary Figure 4. Estimated heritability across HARE and sex strata

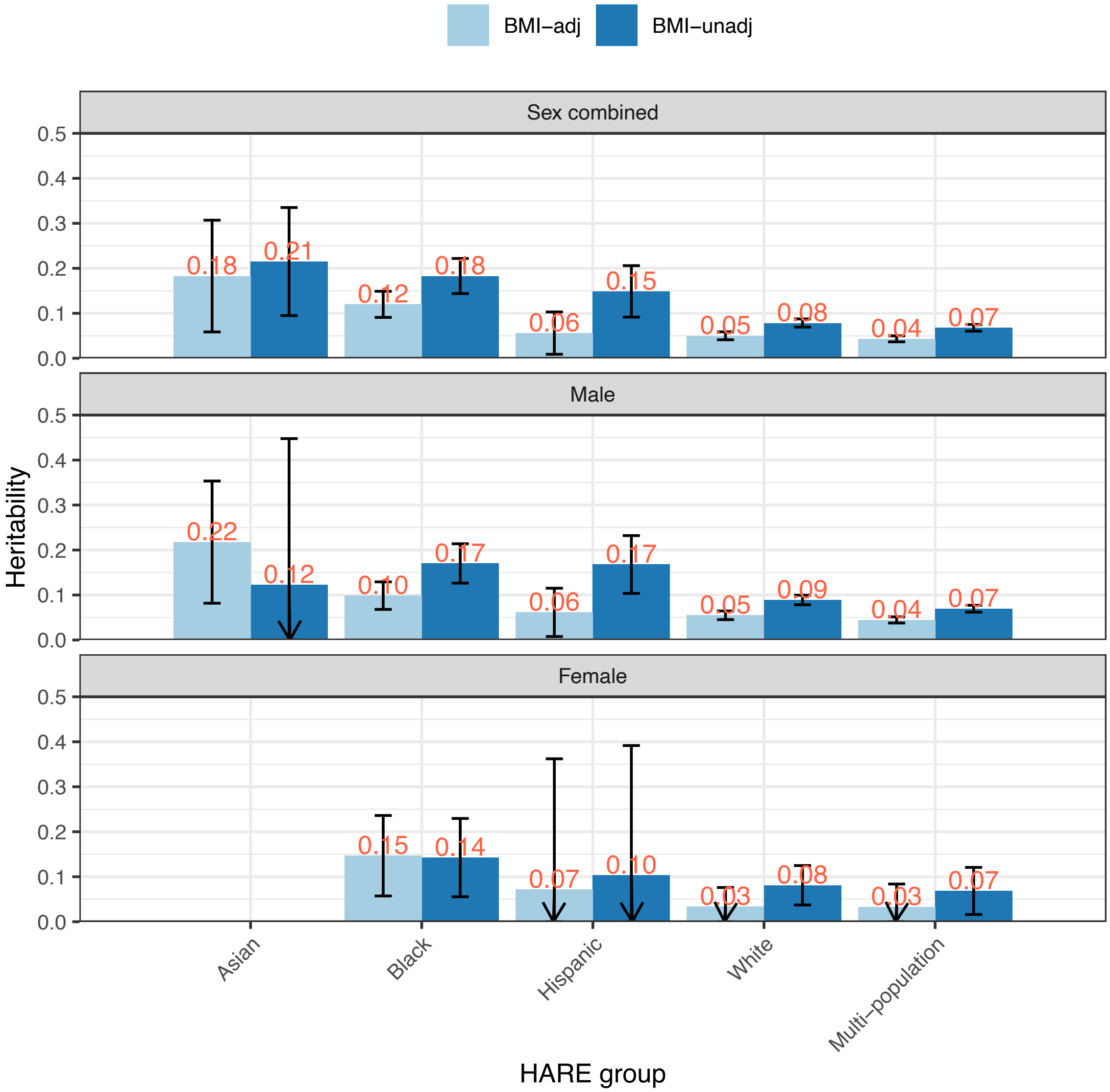

Estimated heritability of BMI-adj and BMI-unadj OSA, HARE and sex stratified and combined analyses. Other than for Asian individuals, we used LDSC to estimate heritability based on summary statistics corresponding to HapMap SNPs. For Asian individuals we used GCTA's implementation of GREML with individual-level data.

#### Supplementary Figure 5. Manhattan and qq-plots of GWAS of sex differences in genetic effects on OSA

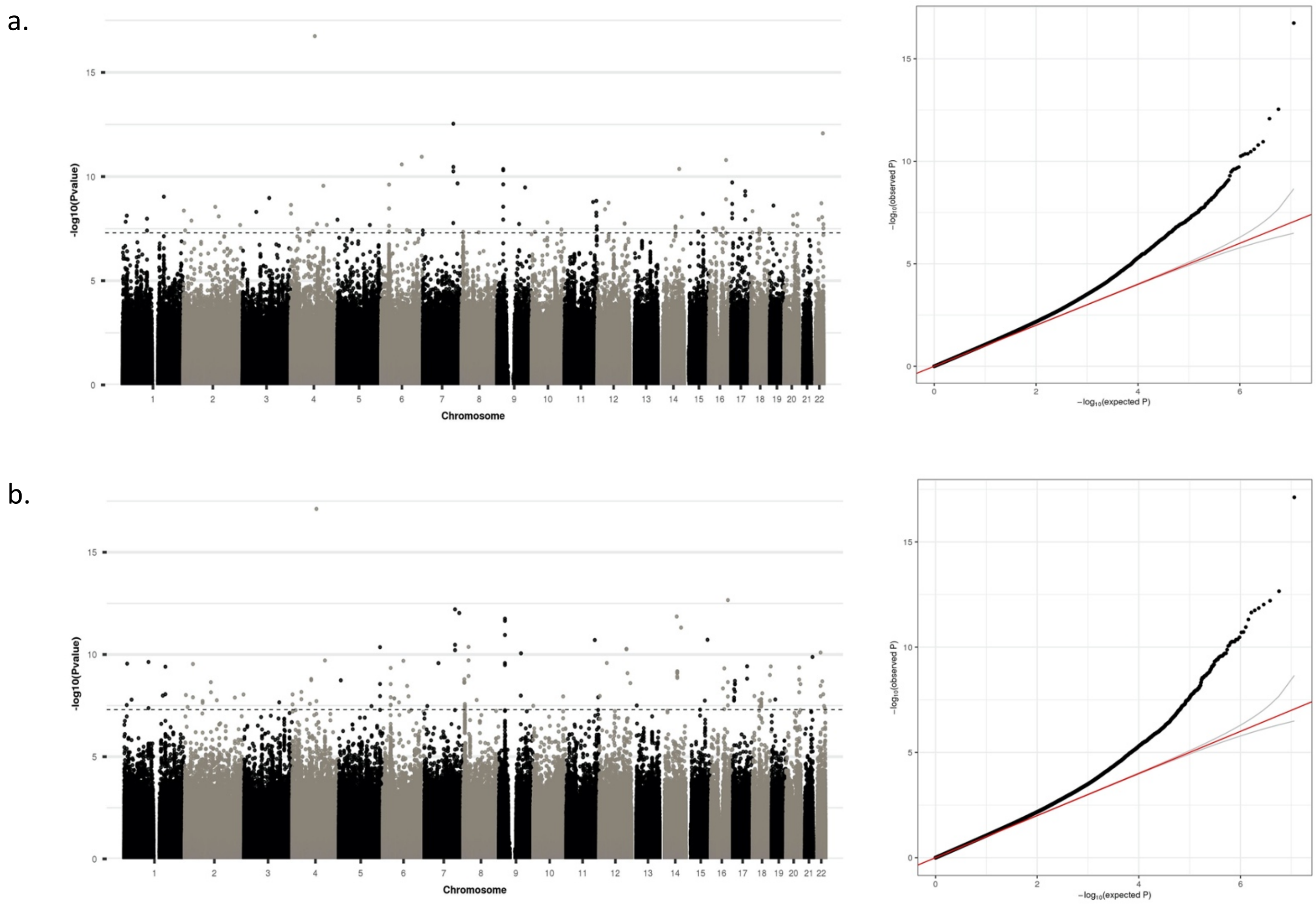

Sex-differences GWAS tested, for each variant, the difference in estimated effect size (log odds ratio) between males and females, while accounting for estimation variance. Panel a provides Manhattan and qq-plots from BMI-unadj analyses, and panel b provides similar results from BMI-adj analyses.

#### Supplementary Figure 6. Comparison of ORs between male and female participants across categories of SNPs, from BMI-unadj analysis

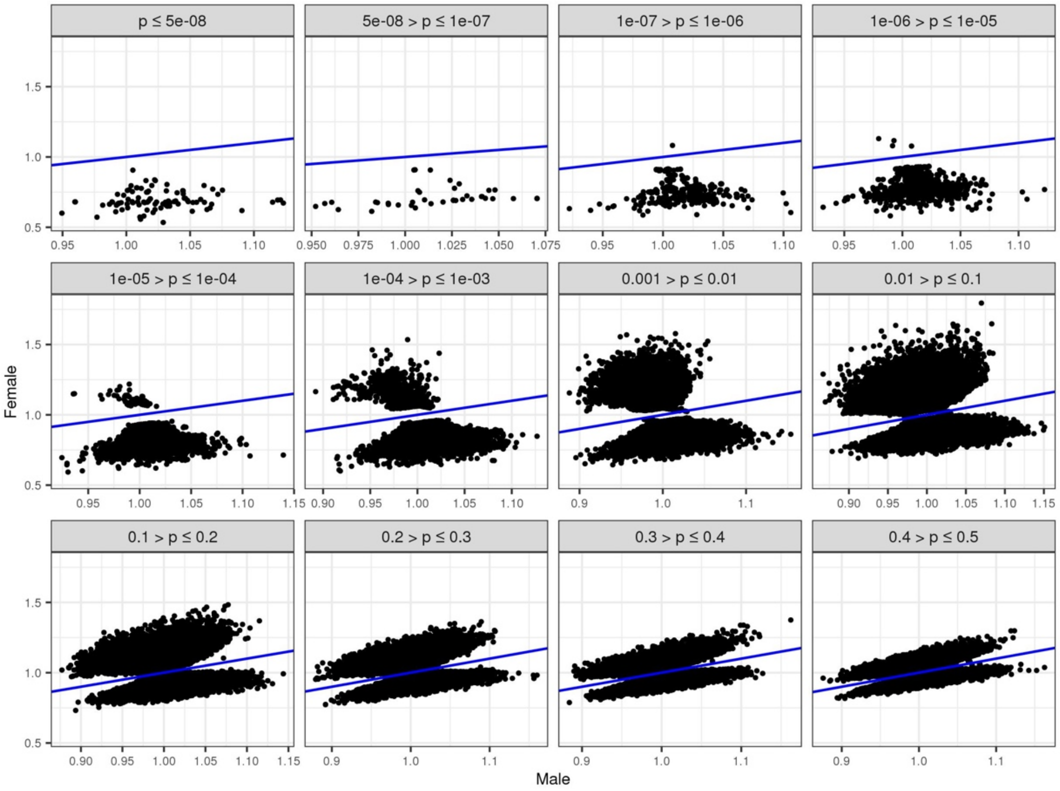

For sets of SNPs defined by their p-value (p) in test of difference in effect sizes between biological sex groups, the figure compares the estimated odds ratio (OR) in males and in females. Depicted results are from BMI-unadj analysis.

#### Supplementary Figure 7. Counts of previously-reported associations of sex-different OSA SNPs by trait group.

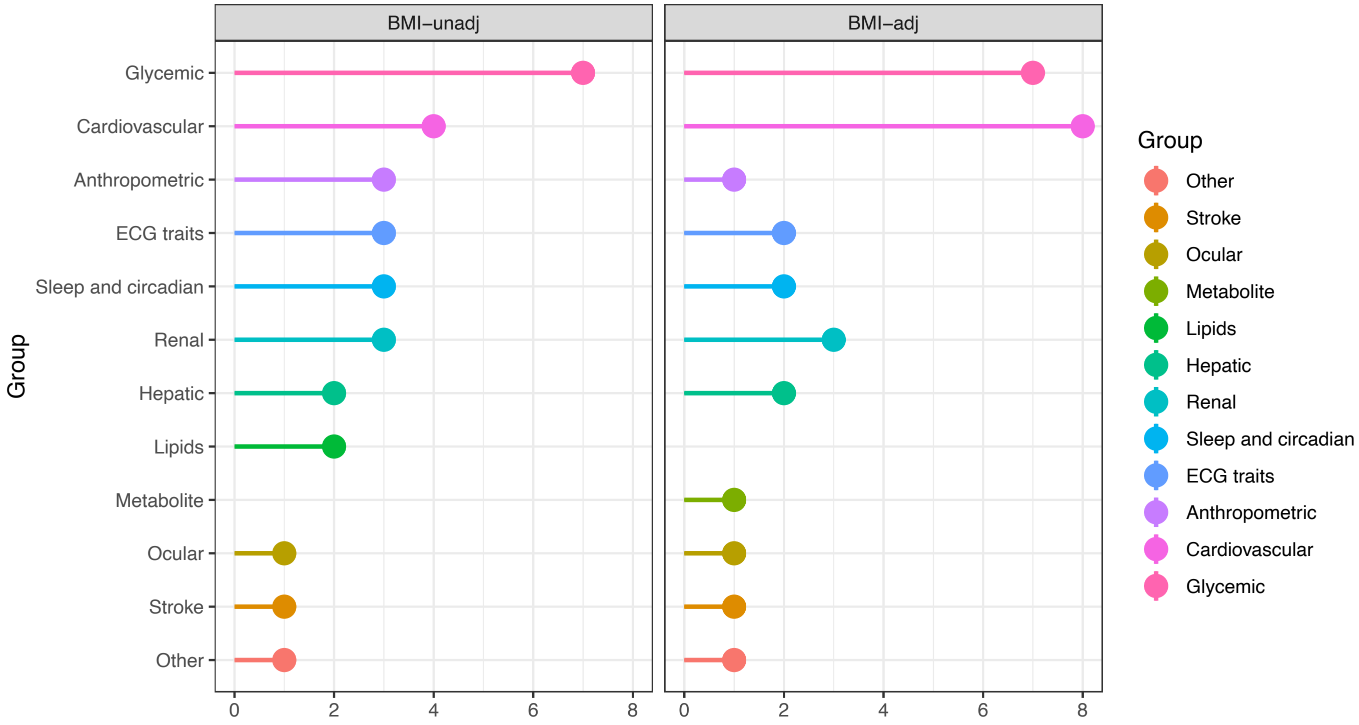

We performed manual searchers at the Type 2 Diabetes Knowledge Portal for each SNPs with genome-wide significant sex-different OSA association. The figure provides the count of SNPs with reported genome-wide significant associations (typically sex-combined) by trait group. Left: counts based on BMI-unadj analysis, right: based on BMI-adj analysis.

Supplementary Figure 8. Manhattan and qq-plots of GWAS of White and Black HARE group differences in genetic effects on OSA.

a.

b.

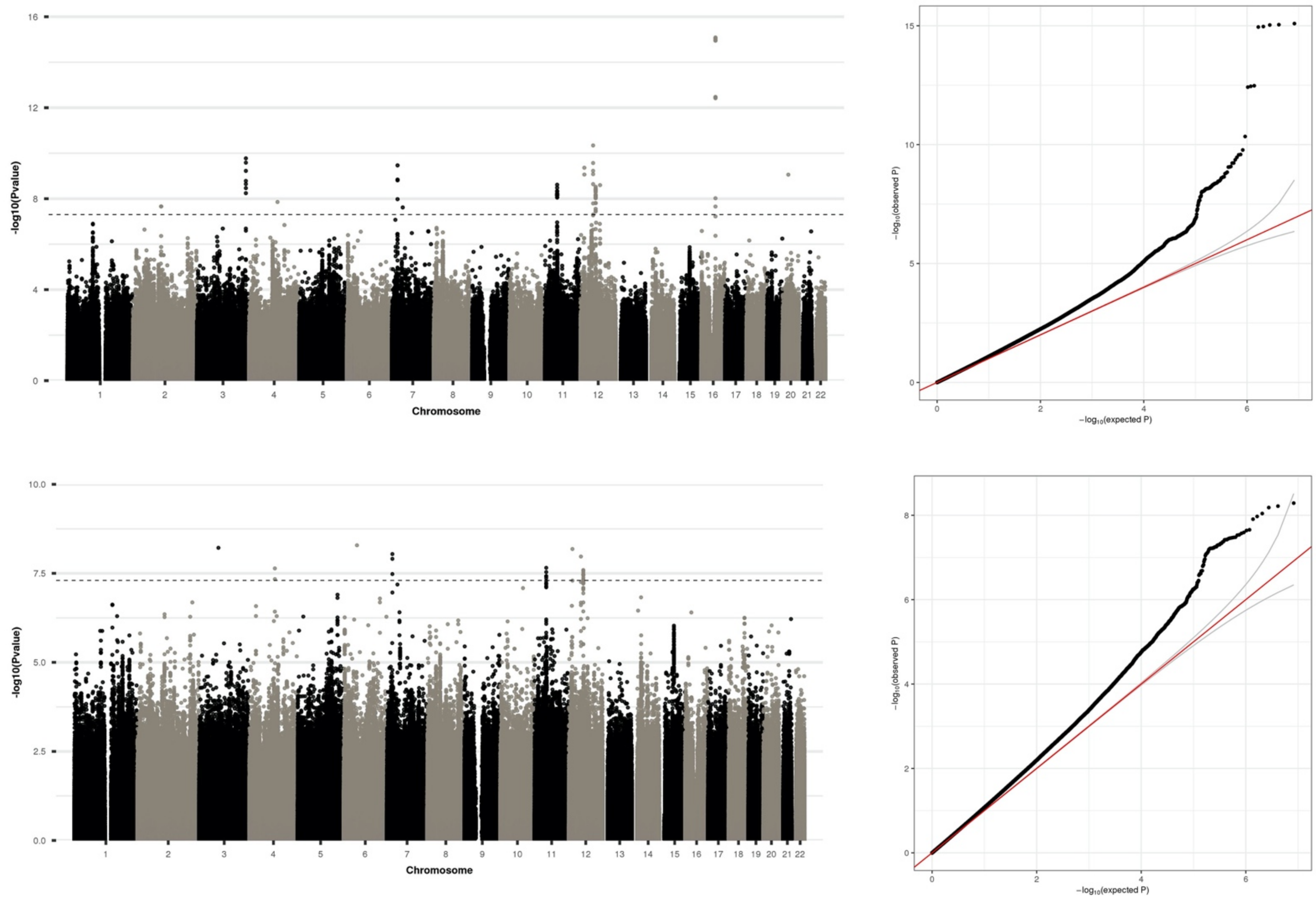

HARE-differences GWAS tested, for each variant, the difference in estimated effect size (log odds ratio) between White and Black HARE groups, while accounting for estimation variance. Panel a provides Manhattan and qq-plots from BMI-unadj analyses, and panel b provides similar results from BMI-adj analyses.

Supplementary Figure 9. Evidence of generalizability of genetic associations between HARE, sex, and obesity groups

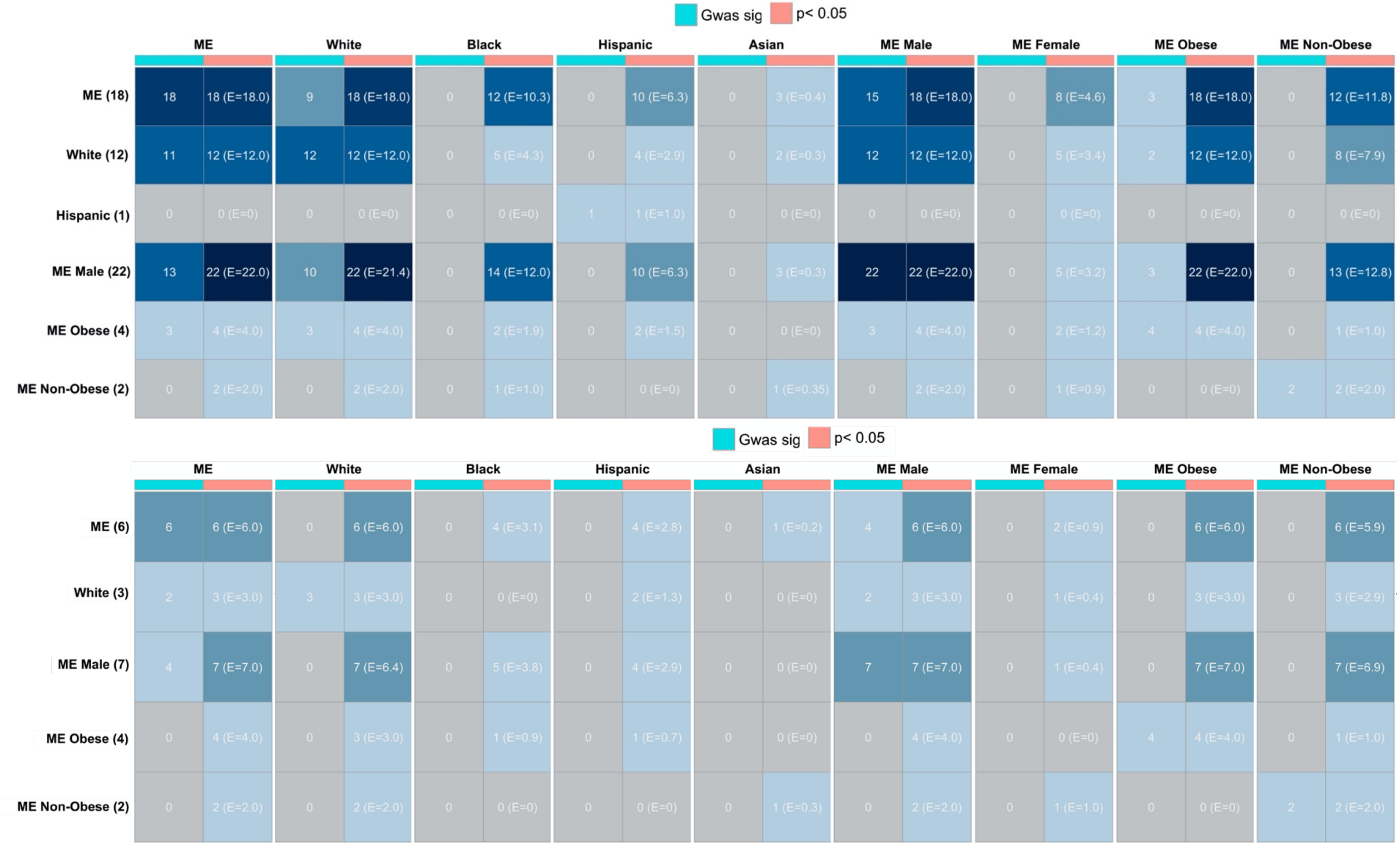

For each of sex, HARE, and obesity strata, as well as the multi-population sex combined meta-analysis, if there were any genome-wide significant associations we looked up whether the associations were detected in other strata at either the genome-wide significance level, or at the p<0.05 level. Each row corresponds to a stratum, and the number in parenthesis next to the row label represents the number of genome-wide significance associations (after clumping) in this stratum. For each of the strata, rows provide the number of SNPs out of those in the row with $p<5x10^{-8}$ ("GWAS sig") and with $p<0.05$. In the columns corresponding to SNP look up based on $p<0.05$, we also provide the expected number of generalization E. The top panel corresponds to BMI-unadj and the bottom to BMI-adj analyses.
